# Long COVID neuropsychological deficits after severe, moderate or mild infection

**DOI:** 10.1101/2021.02.24.21252329

**Authors:** P. Voruz, G. Allali, L. Benzakour, A. Nuber-Champier, M. Thomasson, I. Jacot, J. Pierce, P. Lalive, K-O. Lövblad, O. Braillard, M. Coen, J. Serratrice, J. Pugin, R. Ptak, I. Guessous, B.N. Landis, F. Assal, J.A. Péron

**Affiliations:** Clinical and Experimental Neuropsychology Laboratory, Faculty of Psychology, University of Geneva, Geneva, Switzerland; Neurology Department, Geneva University Hospitals, Switzerland; Faculty of Medicine, University of Geneva, Switzerland; Psychiatry Department, Geneva University Hospitals, Switzerland; Diagnostic and Interventional Neuroradiology Department, Geneva University Hospitals, Switzerland; Division and Department of Primary Care, Geneva University Hospitals, Switzerland; Internal Medicine Department, Geneva University Hospitals, Switzerland; Intensive Care Department, Geneva University Hospitals, Switzerland; Neurorehabilitation Department, Geneva University Hospital, Switzerland; Rhinology-Olfactology Unit, Otorhinolaryngology Department, Geneva University Hospitals, Switzerland

## Abstract

**Background:** There is growing awareness that severe acute respiratory syndrome coronavirus 2 (SARS-CoV-2) infection can include long-term neuropsychological deficits, even in its mild or moderate respiratory forms.

**Methods:** Standardized neuropsychological, psychiatric, neurological and olfactory tests were administered to 45 patients (categorized according to the severity of their respiratory symptoms during the acute phase) 236.51 ± 22.54 days post-discharge following SARS-CoV-2 infection.

**Results:** Deficits were found in all the domains of cognition and the prevalence of psychiatric symptoms was also high in the three groups. The severe performed more poorly on long-term episodic memory and exhibited greater anosognosia. The moderate had poorer emotion recognition, which was positively correlated with persistent olfactory dysfunction. The mild were more stressed, anxious and depressed.

**Conclusion:** The data support the hypothesis that the virus targets the central nervous system (and notably the limbic system), and support the notion of different neuropsychological phenotypes.

## INTRODUCTION

The presence of long-term neuropsychological deficits following severe acute respiratory syndrome coronavirus 2 (SARS-CoV-2) infection is strongly suspected, even in its mild or moderate forms. This is based on four main arguments.

*First*, longitudinal studies of SARS-CoV and the Middle East respiratory syndrome, which share many pathogenetic similarities with SARS-CoV-2, have demonstrated the presence of sleep disorders, frequent recall of traumatic memories, emotional lability, impaired concentration, fatigue, and impaired memory in more than 15% of affected patients 1 month to 3.5 years following infection (Rogers et al., 2020).

*Second*, neurological and cognitive symptoms observed in 38.6% of patients in the acute phase (Mao et al., 2020) are hypothesized to have similar pathophysiological causes to those responsible for short- and long-term cognitive impairment in other pathologies.

Neuropsychological studies among patients with neuro-immunological diseases such as HIV (Wendelken & Valcour, 2012), multiple sclerosis (Piras et al., 2003), and encephalitis (van Sonderen et al., 2016), have reported specific long-term deficits in cognitive functions (e.g., memory, executive or emotional processes) with a neuro-infectious and neuro-immunological pathogenesis. Furthermore, increased prevalence of stroke has been reported in patients with COVID-19 (Merkler et al., 2020; Nannoni, de Groot, Bell, & Markus, 2020), leading to additional short- and long-term neurological and cognitive deficits, depending on the location of the lesion, as described for example by Oxley et al. (2020), who examined five patients aged under 50 years with large-vessel stroke.

*Third,* sudden-onset anosmia is a symptom that has been described extremely frequently by patients following infection with SARS-CoV-2, regardless of the severity of their respiratory symptoms (De Maria, Varese, Dentone, Barisione, & Bassetti, 2020; Lee & Lee, 2020). Researchers have identified sustentacular cells as the potential entry point into the olfactory epithelium (Brann et al., 2020). Unlike olfactory neurons, these cells carry ACE2 receptors (Fodoulian et al., 2020). However, the exact extent to which the olfactory epithelium is affected is still unclear, so it is currently impossible to predict which patients with COVID-19 will develop long-term olfactory disorders (Butowt & Bilinska, 2020). It is not known if and how olfactory neurons are affected by disruption of sustentacular cell function. It is also unclear whether the SARS-CoV-2 infection is confined solely to the olfactory epithelium (Vaira et al., 2020), or whether it follows a neuroinvasive pathway via the cribriform plate. Based on other neuro-olfactory pathologies, it has been suggested that entry through the nose-brain barrier is likely and probably underestimated (Doty, 2008; Landis, Vodicka, & Hummel, 2010). Some authors suggest that the olfactory bulb is damaged following COVID-19 infection (Kandemirli, Altundag, Yildirim, Sanli, & Saatci, 2021; Meinhardt et al., 2021). Interestingly, an ^18^F-FDG PET study among patients with SARS-CoV-2 and anosmia highlighted hypometabolism specifically in the neural substrates of the olfactory circuit, which could indicate an attack on the central nervous system (CNS) (pre-/postcentral gyrus, thalamus/hypothalamus, cerebellum and brainstem) via the olfactory pathway (Guedj et al., 2021; Guedj et al., 2020).

*Fourth*, to our knowledge only one study has so far explored the short-term impact (10-40 days post-hospital discharge) of SARS-CoV-2 infection on cognition using a validated and standardized methodology with face-to-face interviews (Almeria, Cejudo, Sotoca, Deus, & Krupinski, 2020). These authors reported short-term disruption of memory, attention, and executive functions. Unfortunately, they did not explore the impact of the severity of the respiratory symptoms. Hampshire et al. (2020) did consider the influence of severity in their study, but only found a trend toward significance, and used online tests that had not been psychometrically validated. Woo et al. (2020) also addressed the short-term (20-105 days post-infection) impact of SARS-CoV-2 in mild or moderate patients, by administering the Modified Telephone Interview for Cognitive Status, a screening battery that was initially developed for the early detection of dementia. They reported memory and attentional deficits in patients compared with matched controls. These approaches had several potential methodological issues, such as the use of an online survey relying on participants’ unverified self-reports (Hampshire et al., 2020), and the failure to collect information about patients’ clinical history or medical antecedents (Almeria et al., 2020; Woo et al., 2020), which may have induced interindividual variability in the results. Moreover, no study has investigated the long-term effects of infection on the instrumental domains (including visuospatial processing, ideomotor praxis, and language) or emotion recognition. Finally, to our knowledge, the impact of psychiatric factors on the cognitive functioning of patients with SARS-CoV-2 has not been studied thus far. Epidemiological studies have highlighted the impact of the pandemic and related health measures such as lockdown on mental health (Bäuerle et al., 2020; Röhr et al., 2020; Salari et al., 2020), with increased anxiety and depressive symptoms (Wang et al., 2020) within the general population. Being infected by SARS-CoV-2 also has a major affective impact (Almeria et al., 2020). Long-term psychiatric consequences of COVID-19 described so far include anxiety, depressive symptoms, insomnia, and posttraumatic stress disorder (PTSD) (Mazza et al., 2020), especially among patients with a history of psychiatric illness or who required intensive care. All these symptoms may arise from a neurobiological disturbance and the ensuing neuroinflammation process (Kohler, Krogh, Mors, & Eriksen Benros, 2016).

In this context, the present study had three main objectives: i) investigate whether SARS-COV-2 causes long-term (6-9 months after the acute phase) neuropsychological deficits, identify the nature of the affected cognitive and psychiatric domains, and determine their impact on quality of life; ii) explore whether cognitive and psychiatric symptoms are a function of the severity of the respiratory symptoms in the acute phase, and whether patients who present with moderate or even mild forms also exhibit cognitive dysfunctions and/or psychiatric symptoms; and iii) look for correlations between long-term neuropsychological deficits and psychiatric symptoms resulting from a neurobiological disturbance caused by SARS-CoV-2 and/or personal stressful experience in the context of the global health crisis, but also between these deficits and olfactory functions. To this end, patients underwent a comprehensive neuropsychological assessment probing multiple cognitive domains, emotion recognition, psychiatric symptoms, and olfaction. They were divided into three groups according to the respiratory severity of the disease in the acute phase: severe (intensive care with respiratory assistance), moderate (hospitalized without respiratory assistance), or mild (not hospitalized).

Corresponding with our three objectives, we developed three hypotheses. First, we hypothesized that SARS-CoV-2 causes long-term neuropsychological deficits that continue to affect patients’ functioning and quality of life 6-9 months post-infection. We expected to observe cognitive deficits in memory, executive function and logical reasoning (Almeria et al., 2020), as well as the emergence of psychiatric disorders such as anxiety, depressive symptoms, insomnia, and PTSD (Troyer, Kohn, & Hong, 2020; Varatharaj et al., 2020).

Second, we hypothesized that the presence of neuropsychological deficits is positively correlated with disease severity in the acute phase (Hampshire et al., 2020). Third, although ours was an exploratory study, we hypothesized that pandemic- and disease-related psychiatric symptoms explain a significant proportion (but not all) of the variance observed for neuropsychological measures (Baig, Khaleeq, Ali, & Syeda, 2020). Based on Soudry, Lemogne, Malinvaud, Consoli, and Bonfils (2011) and Guedj et al. (2021), we also predicted that a long-term reduction in olfactory performance would correlate positively with any impaired performance on memory and emotion recognition, owing to common neuronal substrates.

## METHODS

### Participants (see Table 1)

Three groups of patients who had been infected with SARS-CoV-2 were included in the study: 15 patients who had been admitted to intensive care during the acute phase of the infection (severe); 15 patients who had been hospitalized but did not require intensive care (moderate); and 15 patients who had tested positive but had not been hospitalized. All the patients had had their infection confirmed by positive polymerase chain reaction (PCR) from nasopharyngeal swab and/or positive serology. On average, the moderate patients had been hospitalized for 9.27 days (± 9.52), and the severe patients for 37.40 days (± 30.50). In comparison with other studies on SARS-CoV-2, the mean duration of hospitalization for the moderate group was somewhat longer, but this was driven by a single patient. The median number of days for this group was 7, which is comparable to that observed in other studies in Switzerland (Regina et al., 2020).

**Table 1:**
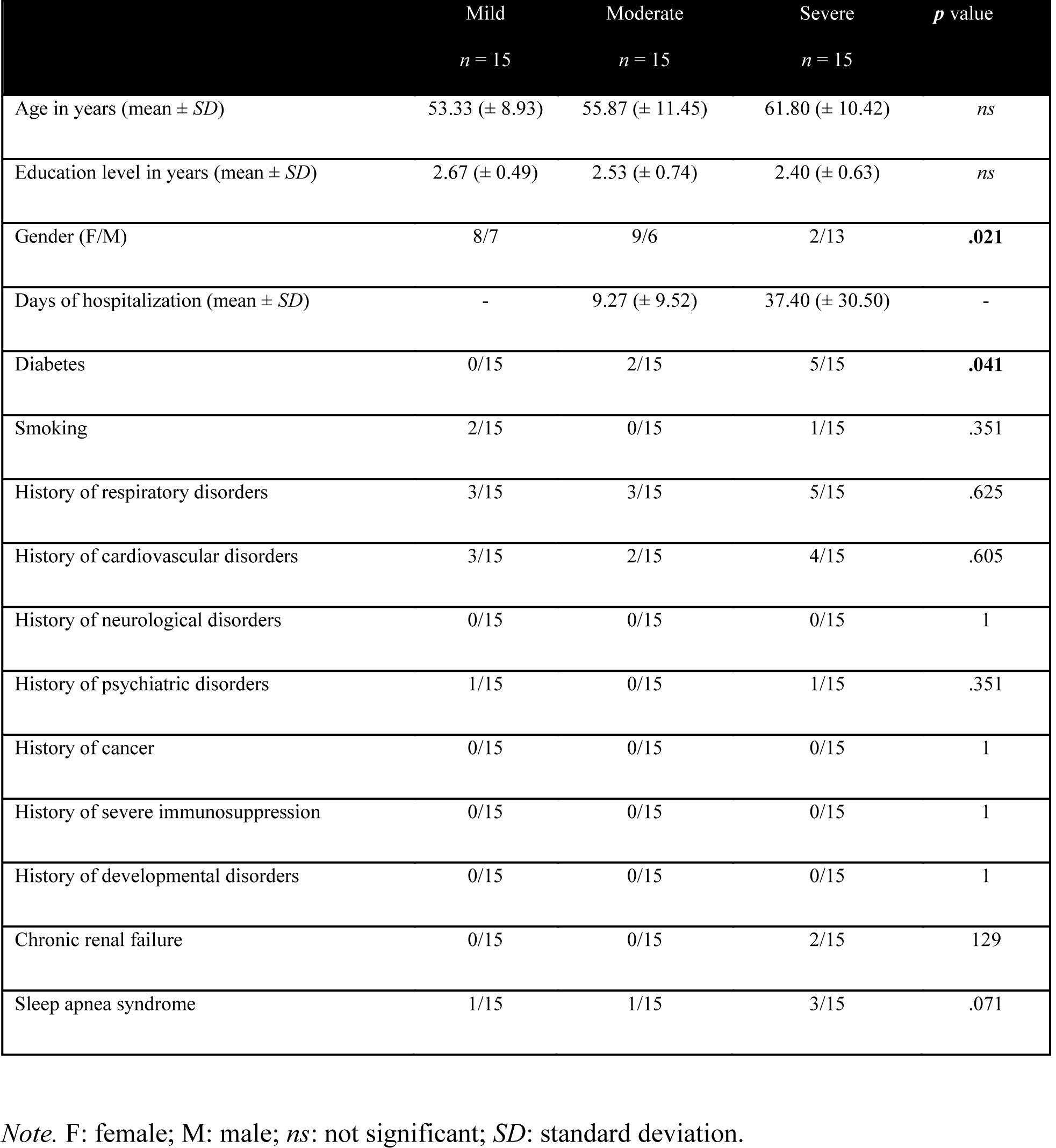
Sociodemographic data and relevant medical antecedents

|  | Mild<br>*n* = 15 | Moderate<br>*n* = 15 | Severe<br>*n* = 15 | *p* value |
| --- | --- | --- | --- | --- |
| Age in years (mean ± *SD*) | 53.33 (± 8.93) | 55.87 (± 11.45) | 61.80 (± 10.42) | *ns* |
| Education level in years (mean ± *SD*) | 2.67 (± 0.49) | 2.53 (± 0.74) | 2.40 (± 0.63) | *ns* |
| Gender (F/M) | 8/7 | 9/6 | 2/13 | **.021** |
| Days of hospitalization (mean ± *SD*) | - | 9.27 (± 9.52) | 37.40 (± 30.50) | - |
| Diabetes | 0/15 | 2/15 | 5/15 | **.041** |
| Smoking | 2/15 | 0/15 | 1/15 | .351 |
| History of respiratory disorders | 3/15 | 3/15 | 5/15 | .625 |
| History of cardiovascular disorders | 3/15 | 2/15 | 4/15 | .605 |
| History of neurological disorders | 0/15 | 0/15 | 0/15 | 1 |
| History of psychiatric disorders | 1/15 | 0/15 | 1/15 | .351 |
| History of cancer | 0/15 | 0/15 | 0/15 | 1 |
| History of severe immunosuppression | 0/15 | 0/15 | 0/15 | 1 |
| History of developmental disorders | 0/15 | 0/15 | 0/15 | 1 |
| Chronic renal failure | 0/15 | 0/15 | 2/15 | 129 |
| Sleep apnea syndrome | 1/15 | 1/15 | 3/15 | .071 |

*Note.* F: female; M: male; *ns*: not significant; *SD*: standard deviation.

The required number of participants in each group was deter e comparison of two means: 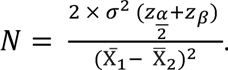. This analysis was based on the literature evaluating the short-term neuropsychological effects of SARS-CoV-2 on mild patients (Woo et al., 2020). To achieve the desired statistical power (1 - β) of 90% and risk of Type I error (α) of 0.05, results indicated that for a one-sided hypothesis, 13 participants were needed in each group. As we planned to perform nonparametric analyses, we had to increase the sample size by 15% (Lehmann, 2012), resulting in 15 participants per group.

The three groups were matched for median age (mild = 57 years; moderate = 55 years; severe = 59 years), sociocultural level, and clinical variables. Given the risk factors associated with the severe form of SARS-CoV-2, there were significantly higher proportions of men (severe = 86.66%; moderate = 40%; mild = 46.66%) and patients with diabetes. Participants were recruited via admission lists provided by the treating doctors at Geneva University Hospitals: LB and OB. For each patient, we carried out a medical file review, followed by a telephone call inviting the patient to take part in the study, if all the eligibility criteria were met. Exclusion criteria were a history of neurological issues, psychiatric disorders (two of the included participants had had an episode of depression more than 10 years before their SARS- CoV-2 infection), cancer (to exclude possible chemotherapy- and radiotherapy-related cognitive impairment (Cascella et al., 2018)), neuro-developmental pathologies, pregnancy, and age above 80 years.

### General procedure and ethics

A flowchart displaying the successive stages of the study according to the eligibility criteria for each experimental group is provided in Figure 1.

**Figure 1:**
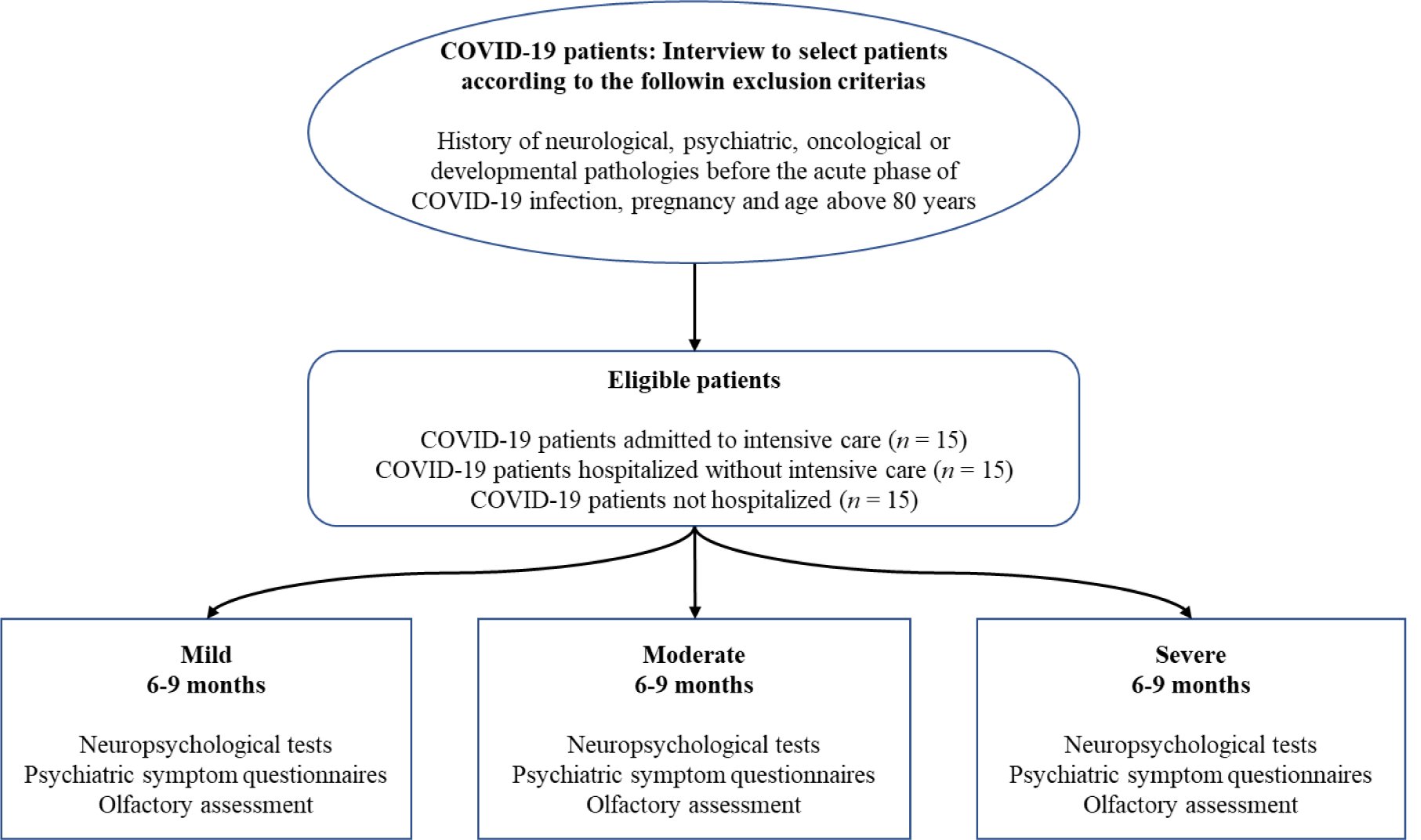
Flowchart of the study

After being given a complete description of the study, participants provided their written informed consent. The study was conducted in accordance with the Declaration of Helsinki, and the study protocol was approved by the cantonal ethics committee of Geneva (CER-02186).

### Neuropsychological assessment

A comprehensive neuropsychological battery was administered to participants 6-9 months after their positive PCR test (236.51 ± 22.54 days). This battery included a series of tests and questionnaires assessing most of the domains of cognition, emotion recognition, fatigue, and quality of life (see below). The tests were administered by clinical psychologists (mean duration: approx. 180 minutes), and the questionnaires were administered online via Qualtrics software (Qualtrics, Provo, UT) (mean duration: approx. 60 minutes).

#### Executive functions

Several tasks were administered to evaluate three executive functions (i.e., inhibition, shifting and updating), in accordance with Miyake et al. (2000): Stroop task, Trail Making Test, and categorical and lexical verbal fluency from the GREFEX battery (Roussel & Godefroy, 2008). Verbal and visuospatial working memory were assessed using the backward digit span (Drozdick, Raiford, Wahlstrom, & Weiss, 2018) and backward Corsi test (Kessels, Van Zandvoort, Postma, Kappelle, & De Haan, 2000). We also administered computer-based tasks designed to gauge focused attention, divided attention, phasic alertness, working memory, and incompatibility using version 2.1 of the Test for Attentional Performance (TAP) (Zimmermann & Fimm, 2007).

#### Memory systems

The short-term memory system was assessed with forward digit spans (Drozdick et al., 2018) and the Corsi test (Kessels et al., 2000). Verbal episodic memory was assessed with the 16-item Grober and Buschke free/cued recall (RL/RI 16) paradigm (Grober & Buschke, 1987), as it distinguishes between the cognitive subprocesses of encoding, storage, and recall (Van der Linden et al., 2004). Visual episodic memory was assessed with the delayed recall of the Rey-Osterrieth Complex Figure test (Meyers & Meyers, 1995).

#### Instrumental function

Language was assessed with the BECLA battery (Macoir, Gauthier, Jean, & Potvin, 2016), ideomotor praxis with a short validated battery (Mahieux- Laurent et al., 2009), visuoconstructive abilities with the Rey-Osterrieth Complex Figure test (Meyers & Meyers, 1995), and visuoperceptual functions with 4 subtests from the Visual Object and Space Perception battery (VOSP) (Warrington & James, 1991) measuring object perception (fragmented letters, object decision) and spatial perception (localization of numbers, analysis of cubes).

#### Logical reasoning

This was assessed using the Puzzle and Matrices subtests of the Wechsler Adult Intelligence Scale–Fourth Edition (WAIS-4) (Wechsler, 2008).

#### Emotion

Multimodal emotion recognition was assessed with the Geneva Emotion Recognition Test (GERT) (Schlegel, Grandjean, & Scherer, 2014). In this emotion recognition task, participants watched 42 video clips, in which 10 actors displayed 14 different emotions (pride, fun, joy, pleasure, relief, interest, anger, irritation, fear, anxiety, disgust, despair, sadness, surprise) while expressing nonverbal content. After each clip, participants were asked to choose one emotion from the list of 14 that best described the emotion played by the actor.

#### Anosognosia and cognitive complaints

We administered the Cognitive Complaints Questionnaire (QPC) (Thomas-Antérion, Ribas, Honoré-Masson, Million, & Laurent, 2004) and the Behavior Rating Inventory of Executive Function-adult Version (BRIEF-A) (Roth, Gioia, & Isquith, 2005). To quantify anosognosia, we calculated a self-appraisal discrepancy (SAD) score for each memory and executive domain evaluated by the QPC and BRIEF-A (Leicht, Berwig, & Gertz, 2010; Rosen et al., 2010; Tondelli et al., 2018). First, we calculated standardized scores for the cognitive complaints, dividing the raw scores of the self-report questionnaires into four categories: 0 = normal behavior; 1 = limited influence on daily life; 2 = noticeable influence on daily life; and 3 = substantial influence on daily life. Then, each standardized score yielded by one of these self-administered questionnaires of cognitive complaints was subtracted from the standardized score for the relevant function. For example, if a patient reported no memory disorders (QPC score = 3) but performed very poorly on Grober and Buschke (RL/RI 16) – delayed free recall (score = 0), he/she would exhibit anosognosia for memory dysfunction: 0 (standardized score on episodic memory test) – 3 (score on self-questionnaire of memory complaints) = -3. SAD scores could therefore range from -3 to 3, and any score below 0 indicated anosognosia.

### Other clinical outcomes

We collected patients’ sociodemographic data and medical history. Psychiatric data (including those concerning current fatigue, insomnia, and somnolence), olfactory abilities, and quality of life at the time of the interview were also collected. Finally, a neurological assessment of CNS and peripheral nervous system functions and walking was carried out by two certified neurologists (FA and GA).

#### Sociodemographic and clinical data

In addition to age, collected during the inclusion interview, we recorded patients’ gender, handedness, and education level. To complement information about previous neurological, psychiatric, and developmental conditions and cancer collected during the inclusion interview, we asked patients about previous cardiovascular disease, respiratory disorders, immunosuppression status, sleep apnea syndrome, diabetes, and smoking. Participants were asked to describe the symptoms they had experienced both during the acute phase of infection and currently (6-9 months post-infection), and the number of days they had spent in hospital, where relevant.

#### Psychiatric data

Depression was assessed with the Beck Depression Inventory-Second edition (BDI-II) (Beck, Steer, & Brown, 1996), anxiety with the State-Trait Anxiety Inventory (STAI-S and STAI-T) (Spielberger, Gorsuch, Lushene, Vagg, & Jacobs, 1993), apathy and its distinct subtypes with the Apathy Motivation Index (AMI) (Ang, Lockwood, Apps, Muhammed, & Husain, 2017), PTSD with the Posttraumatic Stress Disorder Checklist for DSM-5 (PCL-5) (Ashbaugh, Houle-Johnson, Herbert, El-Hage, & Brunet, 2016), manic symptoms with the Goldberg Mania Inventory (Goldberg, 1993), dissociative symptoms in the patient’s daily life with the Dissociative Experience Scale (DES) (Carlson & Putnam, 1986), current stress perception with the Perceived Stress Scale – 14 items (PSS-14) (Lesage, Berjot, & Deschamps, 2012), cognitive reappraisal of an emotional episode and expressive emotional suppression capacities with the Emotion Regulation Questionnaire (ERQ) (Gross & John, 2003) and susceptibility to others’ emotions with the Emotion Contagion Scale (ECS) (Doherty, 1997). Finally, fatigue was assessed with the French version of the Fatigue Impact Scale (Debouverie, Pittion-Vouyovitch, Louis, & Guillemin, 2007), potential sleeping disorders with the Insomnia Severity Index (ISI) (Morin, 1993), and symptoms of sleepiness in daily life with the Epworth Sleepiness Scale (Johns, 1991).

#### Olfaction

Olfactory performance was measured with the Sniffin’ Sticks test battery. This test consists of commercially available pens with 16 common odors, which were each presented for 2 s in front of both nostrils. For each odor, patients had to choose between four descriptors in a multiple-choice task. Participants’ scores ranged from 0 to 16. Based on Kobal et al. (2000), we set three thresholds. Patients with an identification score of 0-7 were considered anosmic, 8-12 hyposmic, and 12-16 normosmic.

#### Quality of life

We administered the SF-36 (Bousquet et al., 1994), which distinguishes between the physical and mental aspects of quality of life.

### Statistical analyses

#### Prevalence of neuropsychological deficits and psychiatric symptoms (Objective 1)

For each neuropsychological test, we first compared patients’ performances with normative data for the validated neuropsychological tools. As the standardization depended upon the distribution of the normative data collected from the reference sample (*t* and *z* scores, percentiles, or standard scores), the comparative tests were adjusted according to the guidelines provided by the authors of the validation study for each test. Second, the data were normalized according to the guidelines of the Swiss Association of Neuropsychology (Frei et al., 2016; Heaton, Grant, & Matthews, 1991), making it possible to classify the standardized as follows: far below the norm (< 2^nd^ percentile), substandard (2^nd^-5^th^ percentiles), borderline or below the normal limit (6^th^-15^th^ percentiles), normal (≥ 16^th^ percentile). This standardization allowed us to quantify the prevalence of each type of disorder, while controlling for variables such as age, education level, and gender. To consider the possible effect of fatigue and increase the robustness of the results, only those performances that were far below the norm (< 2^nd^ percentile) or substandard (2^nd^-5^th^ percentile) were used to calculate the prevalence of neuropsychological deficits. Results that were just below the norm were therefore not considered in the prevalence table.

#### Neuropsychological deficits as a function of disease severity (Objective 2)

For each neuropsychological, psychiatric or quality-of-life measure, we compared the three groups (severe, moderate, and mild) on the raw data. Given the distribution of the samples, we used nonparametric Kruskal-Wallis tests. For significant (*p* < .05) measures, Mann-Whitney tests were performed for the 2 x 2 comparisons with Bonferroni-corrected *p* values.

#### Relationships between neuropsychological deficits, psychiatric symptoms, and other secondary variables (Objective 3)

For each neuropsychological variable of interest, forward stepwise multiple regression analyses were performed on the raw cognitive data with the significant sociodemographic variables, sniff test results and psychiatric measures, to quantify relationships between these variables and the neuropsychological functions.

In parallel, and in order to elucidate the cognitive data’s underlying structure, we performed a principal component analysis (PCA) on the raw test and questionnaire scores assessing cognition and emotion recognition. The list of variables included in the PCA is available in Supplementary Information 1. We extracted the first three components with the highest eigenvalues. We then reran forward stepwise multiple regressions for each cognitive component, with the same variables of interest as those described above.

## RESULTS

### Neuropsychological, psychiatric and olfactory profiles 6-9 months post-infection

The first aim of this study was to assess the prevalence of neuropsychological impairments and psychiatric symptoms 6-9 months after SARS-CoV-2 infection. We compared patients’ performances with available normative data to identify the number of impaired scores per patient, group, and test. The prevalence of cognitive impairments in each group 236.51 ± days after infection is set out in Table 2, and the prevalence of psychiatric symptoms in Table 3

**Table 2:**
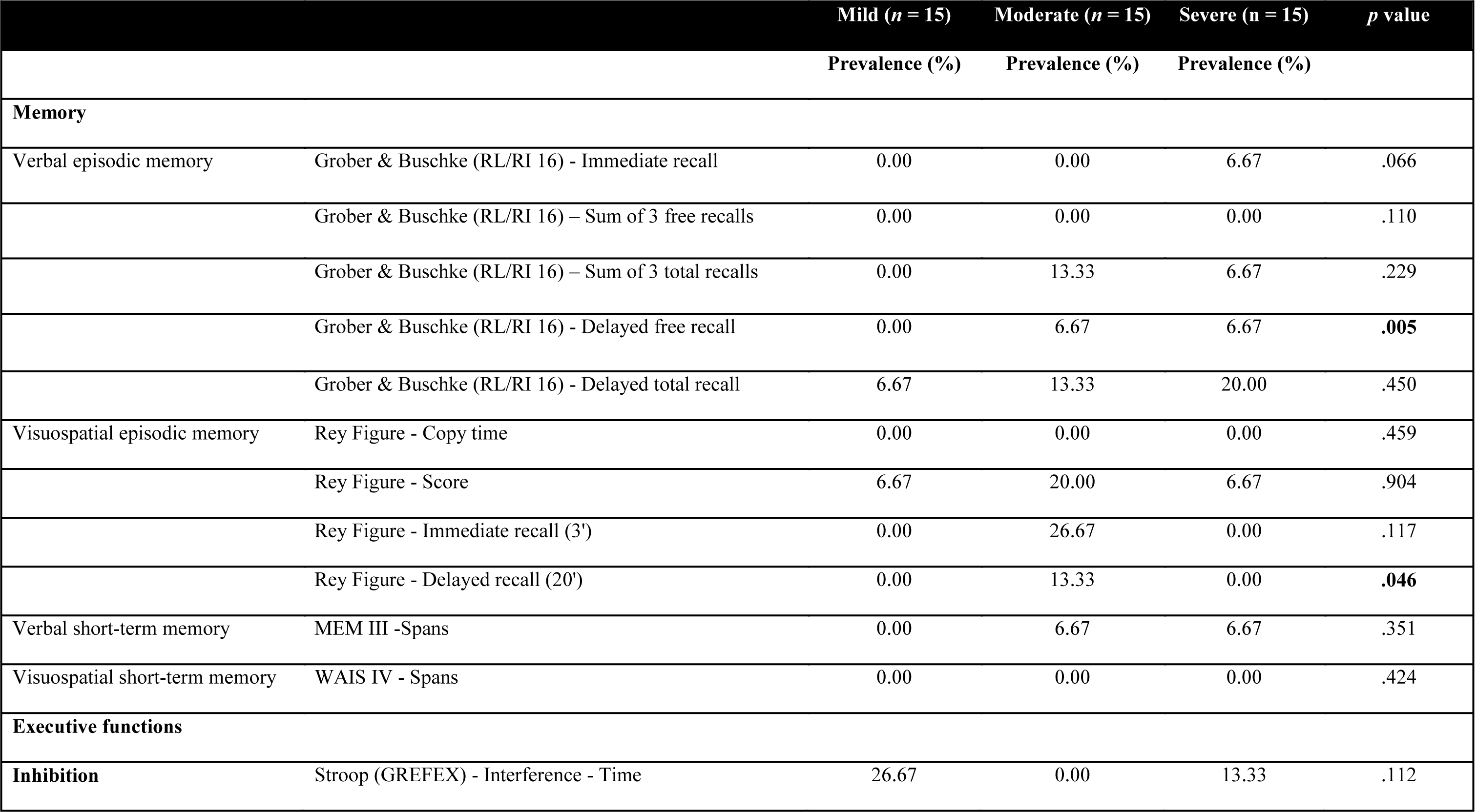
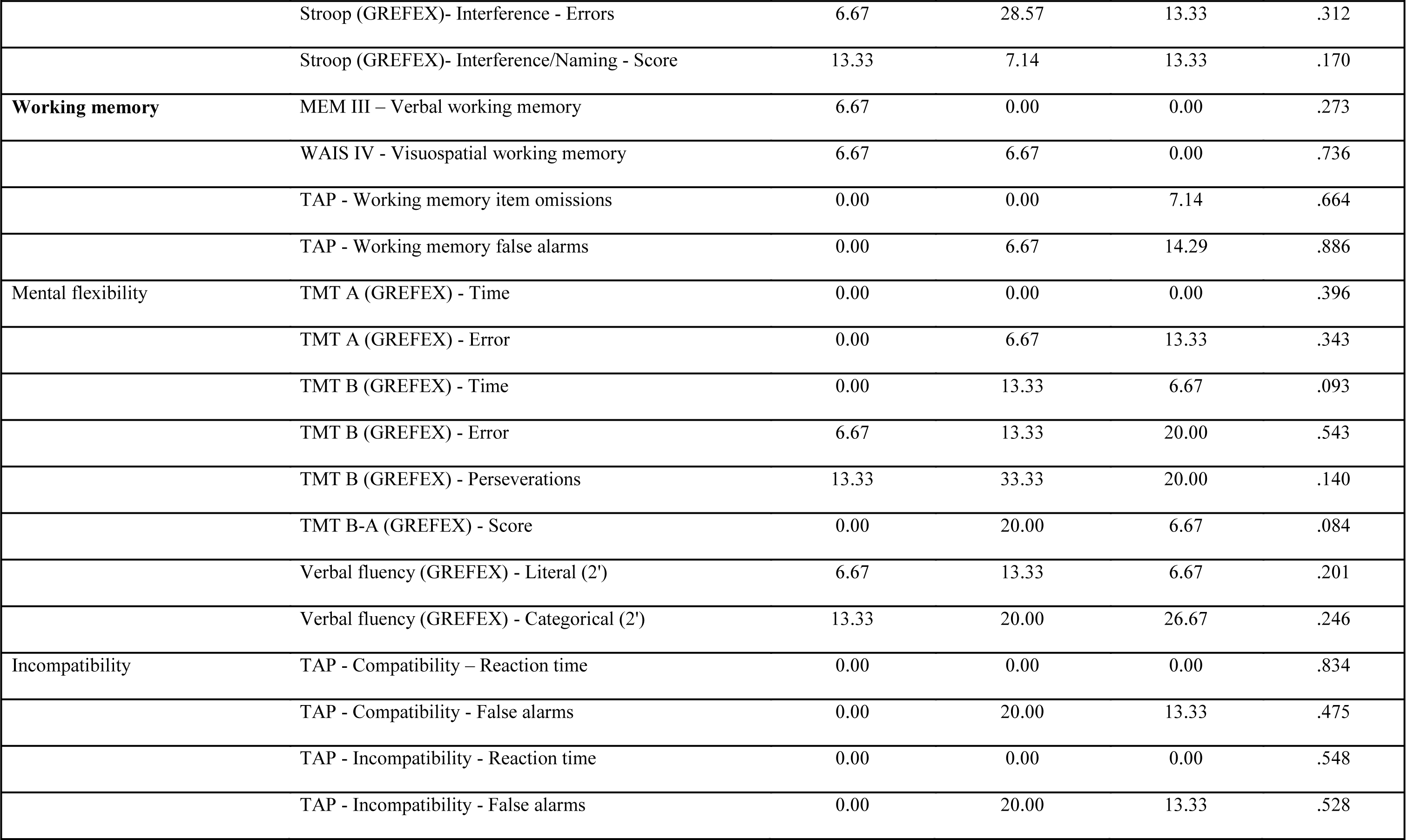
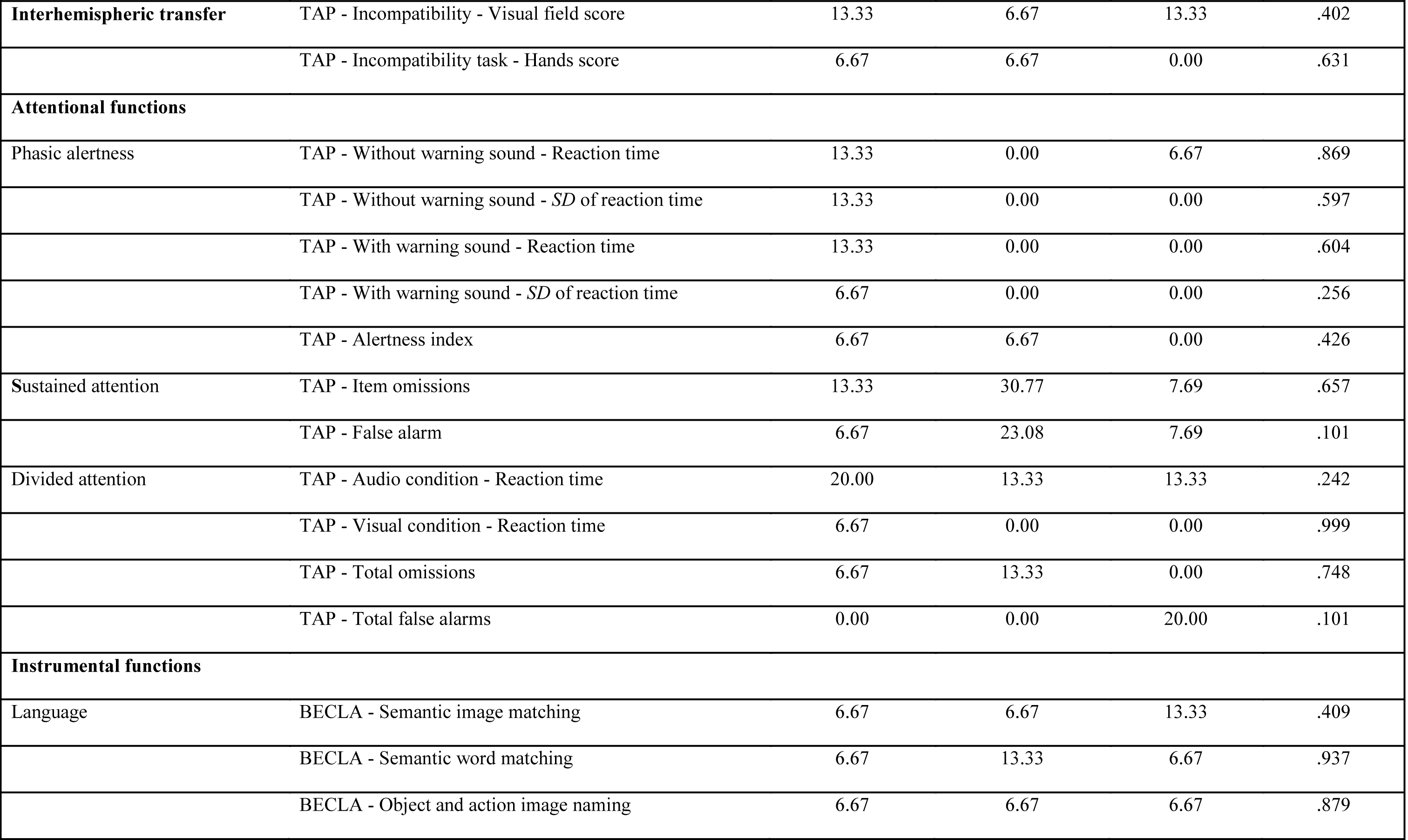
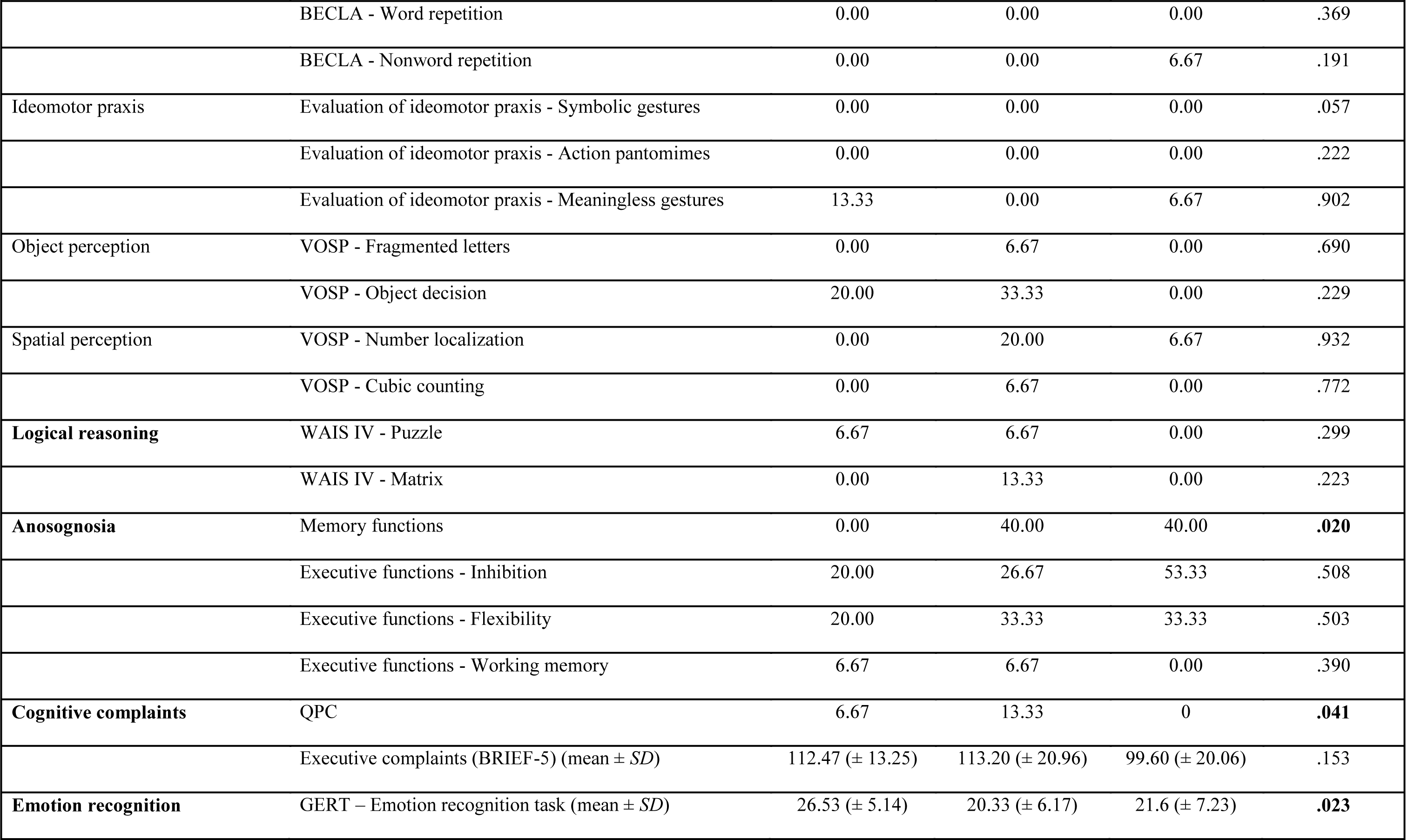
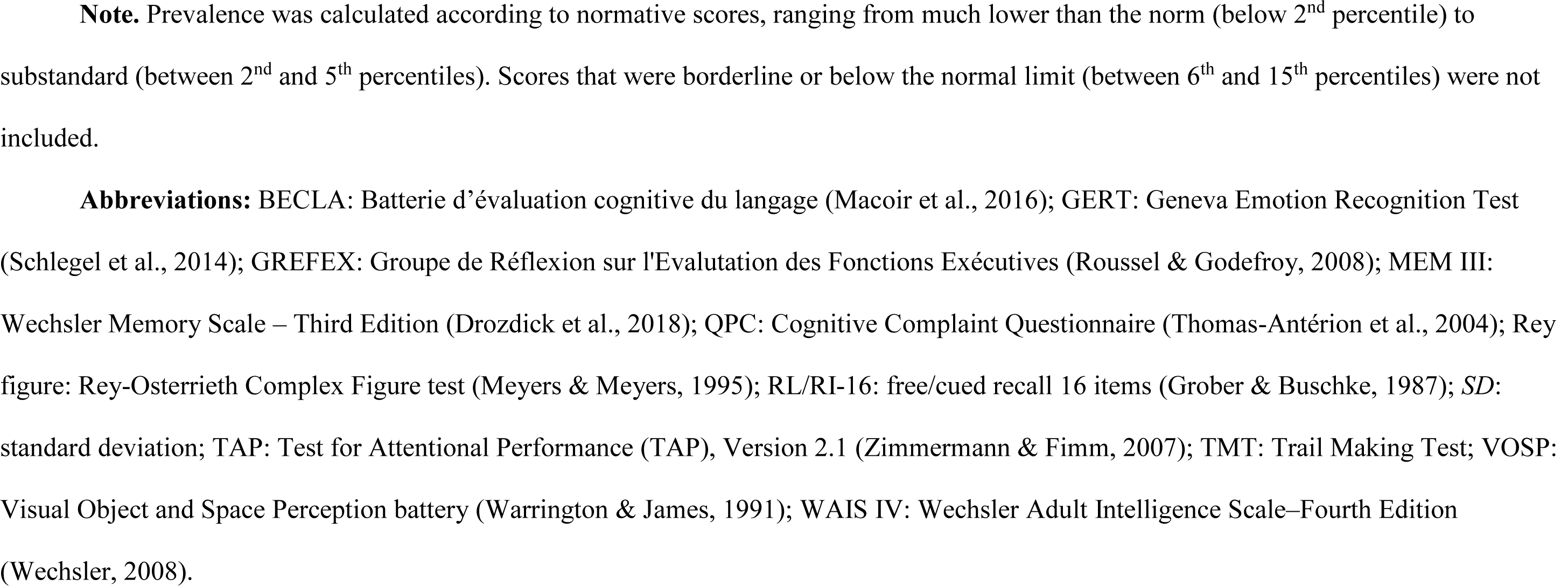
Cognitive deficits among patients with mild, moderate or severe COVID-19 6-9 months post-infection

**Table 3:** Psychiatric symptoms and olfaction in patients with mild, moderate or severe COVID-19 6-9 months post-infection

| Psychiatric symptoms | Mild (*n* = 15) | Moderate (*n* = 15) | Severe (*n* = 15) | *p* value |
| --- | --- | --- | --- | --- |
| Depression (BDI-II) | Minor = 46.67% | Minor = 66.67% | Minor = 80% | **.009** |
| (prevalence) | Mild = 20% | Mild = 20% | Mild = 20% |  |
|  | Moderate = 33.33% | Moderate = 13.33% | Moderate = 0% |  |
|  | Severe = 0% | Severe = 0% | Severe = 0% |  |
| State anxiety (STAI-state) | Very low = 26.67% | Very low = 60% | Very low = 86.67% | **.002** |
| (prevalence) | Low = 33.33% | Low = 6.67% | Low = 6.67% |  |
|  | Moderate = 13.33% | Moderate = 20% | Moderate = 6.67% |  |
|  | High = 26.67% | High = 13.33% | High = 0% |  |
|  | Very high = 0% | Very high = 0% | Very high = 0% |  |
| Trait anxiety (STAI-trait) | Very low = 46.67% | Very low = 73.33% | Very low = 60% | .100 |
| (prevalence) | Low = 26.67% | Low = 0% | Low = 33.33% |  |
|  | Moderate = 13.33% | Moderate = 20% | Moderate = 6.67% |  |
|  | High = 13.33% | High = 6.67% | High = 0% |  |
|  | Very high = 0% | Very high = 0% | Very high = 0% |  |
| Mania (Goldberg Inventory)<br>(prevalence) | Probably absent = 26.67%<br>Hypomania = 26.67%<br>Close to mania = 20%<br>Moderate = 26.67%<br>Ordinary to severe = 0%<br>Severe = 0 | Probably absent = 13.33%<br>Hypomania = 26.67%<br>Close to mania = 20%<br>Moderate = 40%<br>Ordinary to severe = 0%<br>Severe = 0 | Probably absent = 20%<br>Hypomania = 46.67%<br>Close to mania = 6.67%<br>Moderate = 26.67%<br>Ordinary to severe = 0%<br>Severe = 0 | .909 |
| Apathy (AMI-total)<br>(prevalence) | Absent = 86.67%<br>Moderate = 13.33%<br>High = 0% | Absent = 93.33%<br>Moderate = 6.67%<br>High = 0% | Absent = 73.33%<br>Moderate = 26.67%<br>High = 0% | .602 |
| Behavioral apathy (AMI-behavioral)<br>(prevalence) | Absent = 100%<br>Moderate = 0%<br>High = 0% | Absent = 100%<br>Moderate = 0%<br>High = 0% | Absent = 93.33%<br>Moderate = 6.67%<br>High = 0% | .211 |
| Social apathy (AMI-social)<br>(prevalence) | Absent = 93.33%<br>Moderate = 6.67%<br>High = 0% | Absent = 86.67%<br>Moderate = 6.67%<br>High = 6.67% | Absent = 73.33%<br>Moderate = 26.67%<br>High = 0% | .940 |
| Emotional apathy (AMI-emotional)<br>(prevalence) | Absent = 73.33%<br>Moderate = 26.67%<br>High = 0% | Absent = 60%<br>Moderate = 40%<br>High = 0% | Absent = 40%<br>Moderate = 33.33%<br>High = 26.67% | **.029** |
| Posttraumatic stress disorder (PCL-5)<br>(prevalence) | Absent = 86.67%<br>Present = 13.33% | Absent = 86.67%<br>Present = 13.33% | Absent = 93.33%<br>Present = 6.67% | .054 |
| Stress (PSS-14)<br>Mean (± *SD*) | 26.13 (± 9.53) | 19.6 (± 7.47) | 14.93 (± 9.42) | **.023** |
| Dissociative disorder (DES)<br>Mean (± *SD*) | 7.68 (± 11.89) | 10.45 (± 9.23) | 3.98 (± 3.03) | .140 |
| Emotional contagion (ECS)<br>Mean (± *SD*) | 41.40 (± 7.20) | 44.6 (± 4.22) | 36.13 (± 8.38) | **.002** |
| Emotional regulation (ERQ)<br>Mean (± *SD*) | 41.6 (± 7.39) | 44.2 (± 8.33) | 39.80 (± 11.77) | .416 |
| Somnolence (Epworth)<br>(prevalence) | Pathological = 40.00% | Pathological = 53.33% | Pathological = 26.67% | **.036** |
| Insomnia (ISI)<br>(prevalence) | Absent = 20%<br>Mild = 40%<br>Moderate = 40%<br>Severe = 0% | Absent = 46.67%<br>Mild = 40%<br>Moderate = 13.33%<br>Severe = 0% | Absent = 60%<br>Mild = 26.67%<br>Moderate = 13.33%<br>Severe = 0% | **.040** |
| Fatigue (EMIF-SEP)<br>(prevalence) | Present = 13.33% | Present = 20% | Present = 6.67% | .088 |
| Sniff test (anosmia)<br>Mean (± *SD*) | 13.07 (± 1.44) | 11.53 (± 2.13) | 11.47 (± 2.90) | .067 |

**Abbreviations:** AMI-behavioral: Apathy Motivation Index – behavioral score (Ang et al., 2017); AMI-emotional: Apathy Motivation Index – emotional score (Ang et al., 2017); AMI-social: Apathy Motivation Index – social score (Ang et al., 2017); AMI-total: Apathy Motivation Index – total score (Ang et al., 2017); BDI-II: Beck Depression Inventory-Second edition (Beck et al., 1996); DES: Dissociative Experience Scale (Carlson & Putnam, 1986); ECS: Emotion Contagion Scale (Doherty, 1997); EMIF-SEP: Fatigue Impact Scale, French adaptation (Debouverie et al., 2007); ERQ: Emotion Regulation Questionnaire (Gross & John, 2003); Epworth: Epworth Sleepiness Scale (Johns, 1991); Goldberg Inventory: Goldberg Mania Inventory (Goldberg, 1993); ISI: Insomnia Severity Index (Morin, 1993); PCL-5: Posttraumatic Stress Disorder Checklist for DSM-5 (Ashbaugh et al., 2016); PSS-14: Perceived Stress Scale – 14 items (Lesage et al., 2012); STAI-trait: State- Trait Anxiety Inventory (Spielberger et al., 1993); STAI-state: State-Trait Anxiety Inventory (Spielberger et al., 1993).

#### Cognition

Cognitive deficits common to all three groups were observed in the following domains: long-term episodic memory in both the verbal and visual modalities, executive functions (e.g., inhibition and mental flexibility, and both categorical and literal verbal fluency), sustained and divided attention, and language (semantic matching and naming). All three groups exhibited anosognosia for executive dysfunction (see Table 2)

#### Psychiatric disorders

All three groups displayed anxiety, mania, the social component of apathy, stress, PTSD, and dissociative disorders. All three groups also reported insomnia, fatigue and pathological somnolence (see Table 3). The only psychiatric variable where the prevalence score stood out for severe patients was emotional apathy, as measured with AMI (see Table 3).

#### Olfaction

33.33% of the mild group, 73.33% of the moderate group, and 46.66% of the severe group displayed hyposmia. There was no anosmia in the mild and moderate groups, but 13.33% of the severe group were anosmic (see Table 3).

### Neuropsychological and psychiatric symptoms as a function of disease severity

The second aim was to determine whether cognitive deficits and psychiatric symptoms are a function of the severity of the respiratory symptoms in the acute phase. To this end, we compared the three groups on neuropsychological, psychiatric and other clinical data (Kruskal-Wallis statistics and *p* values reported in Tables 2, 3 and 4; Bonferroni-corrected Mann-Whitney statistics and *p* values reported below).

**Table 4:**
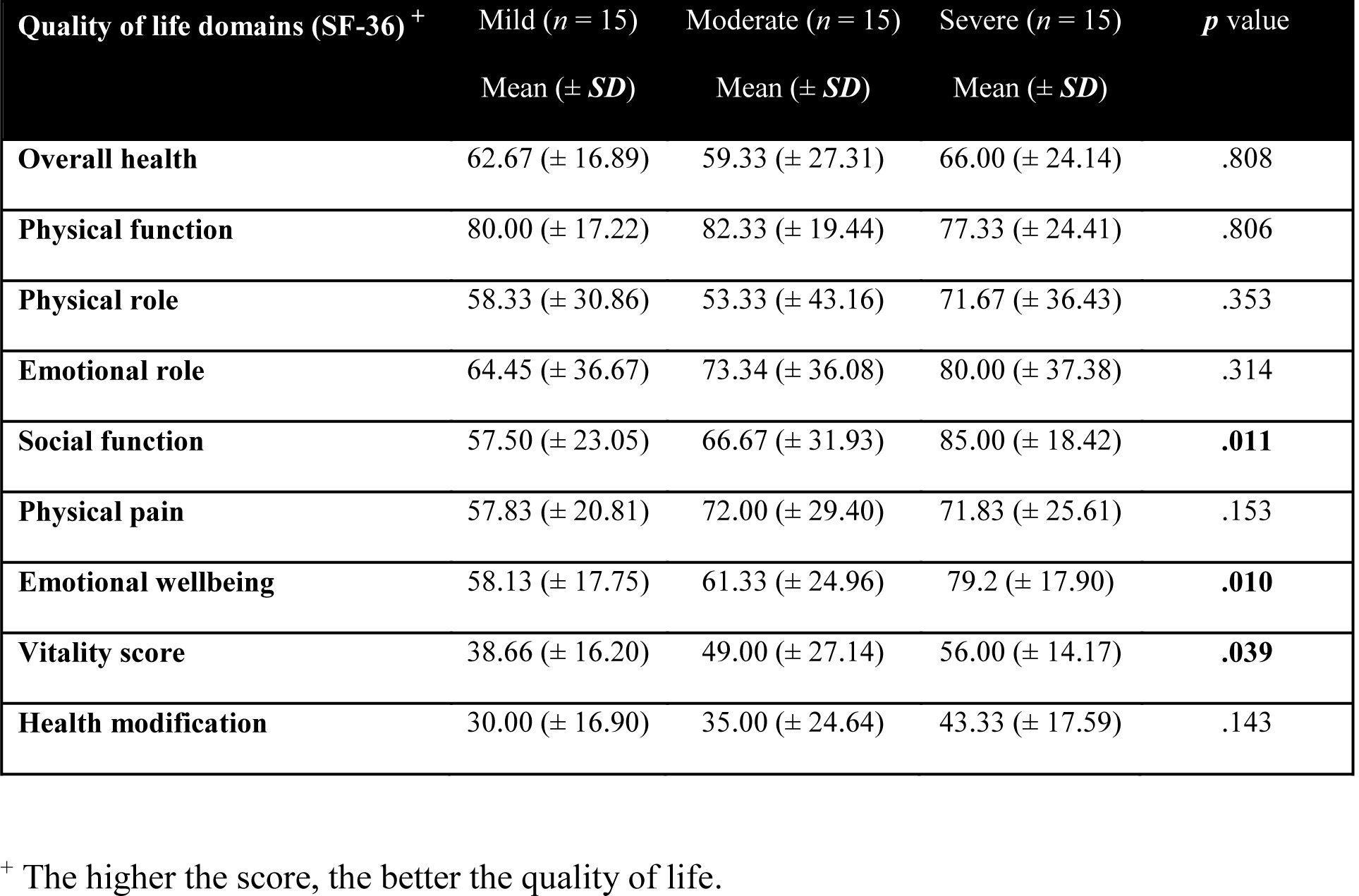
Quality of life of patients with mild, moderate or severe COVID-19 6-9 months post-infection

#### i) Neuropsychological data (Fig. 2)

The three groups differed significantly on i) long-term episodic memory in both the verbal (Grober and Buschke (RL/RI 16) delayed free recall, *H* = 10.75, *p* = .005) and visual (Rey Figure delayed free recall, *H* = 6.15, *p* = .046) modalities, ii) multimodal emotion recognition (GERT; *H* = 7.55, *p* = .023), iii) cognitive complaints (QPC; *H* = 6.38, *p* = .041) and anosognosia for memory dysfunction (SAD; *H* = 7.84, *p* = .020). The other effects were not significant (*p* > .05 for all comparisons).

**Figure 2.**
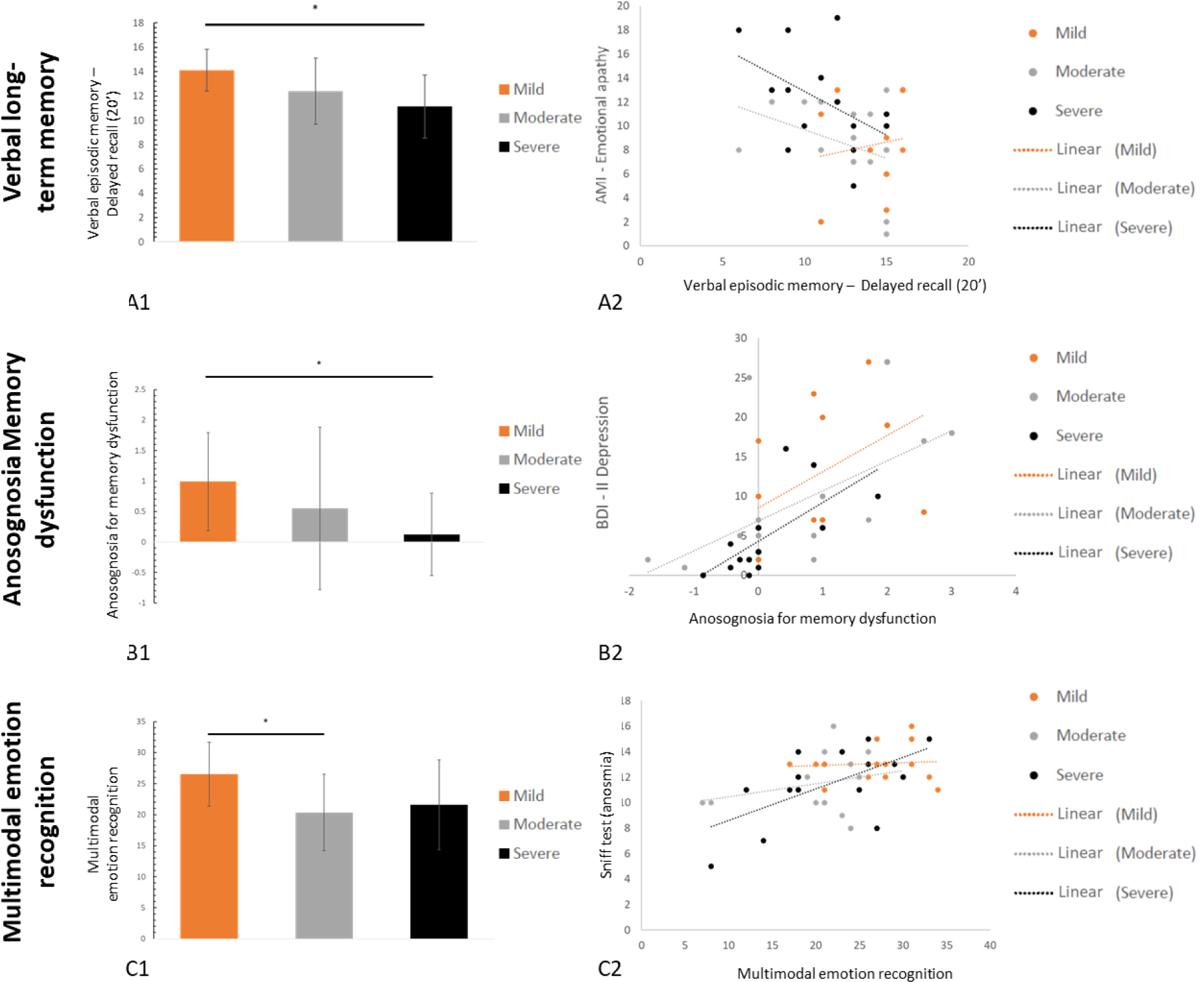
Mean ratings (and standard deviations) for all three groups (severe in black, moderate in gray, and mild in orange) on tasks evaluating verbal episodic memory (A1), anosognosia for memory dysfunction (B1), and multimodal emotion recognition (C1), as well as their respective predictors (A2, B2 and C2) **Note.** A2: The greater the emotional apathy, the poorer the performance on verbal memory (except for the mild group); B2: The lower the depression, the greater the anosognosia for memory dysfunction; C2: The poorer the olfactory recognition, the poorer the emotion recognition.

##### Episodic memor

For Grober and Buschke delayed free recall, the mild patients scored significantly higher than the severe patients (*z*= 3.04, *p* = .002), but the other two pairwise comparisons were not significant after Bonferroni correction (moderate vs. severe: *z* = -1.47, *p* = .141; mild vs. moderate: *z* = 2.00, *p* = .046). Pairwise comparisons were not significant for visual episodic memory (mild vs. moderate: *z* = 2.26, *p* = .023; mild vs. severe: *z* = 0.48, *p* = .61; moderate vs. severe: *z* = 1.89, *p* = .059).

##### Emotion recognition

Mild patients scored significantly higher than moderate patients (*z* = 2.61, *p* = 0.009), but neither the difference between mild and severe patients (*z* = 1.97, *p* = 0.048), nor the difference between moderate and severe patients (*z* = .49, *p* = .620) reached significance after Bonferroni correction.

##### Cognitive complaints and anosognos

Mild patients had more cognitive complaints than severe patients (*z* = -2.55, *p* = .010), but there were no differences between either the mild and moderate patients (*z* = -1.31, *p* = .191), or the moderate and severe patients (*z* = - 0.93, *p* = .351). By contrast, severe patients exhibited more anosognosia for memory dysfunction than mild patients did (*z* = 2.97, *p* = .003), while there were no differences between either the mild and moderate patients (*z* = 1.41, *p* = .158) or the moderate and severe patients (*z* = - 0.76, *p* = .443).

#### ii) Psychiatric data

The three groups differed significantly on depression (*H* = 9.40, *p* = .009), state anxiety (*H* = 12.93, *p* = .002), emotional apathy (*H* = 7.10, *p* = .029), stress (*H* = 7.55, *p* = .023), and emotional contagion (*H* = 9.73, *p* = .002). The other effects were not significant (*p* > .05 for all comparisons). Pairwise comparisons for each of these group differences are described below.

##### Depression, stress, and state anxiety

The mild patients were more depressed, stressed and anxious than the severe patients (BDI-II: *z* = -2.99, *p* = .003; PSS: *z* = -2.55, *p* = .010; STAI-S: *z* = -3.57, *p* < .001), while there were no differences between either the severe and moderate patients (BDI-II: *z* = -1.38, *p* = .165; PSS: *z* = -1.08, *p* = .281; STAI-S: *z* = -1.76, *p* = .078) or the mild and moderate patients (BDI II: *z* = -1.66, *p* = .097; PSS: *z* = -1.08, *p* = .281; STAI-S: *z* = -1.72, *p* = .085).

##### Apathy

For the AMI emotional subscore, pairwise comparisons failed to reach significance after Bonferroni correction (severe vs. mild: *z* = 2.32, *p* = .020; severe vs. moderate: *z* = 2.20, *p* = .028, mild vs. moderate: *z* = 0.08, *p* = .933).

##### Emotional contagion

Pairwise comparisons failed to reach significance after Bonferroni correction (severe vs. moderate: *z* = -3.03, *p* = .017; severe vs. mild: *z* = -1.89, *p* = .059; moderate vs. mild: *z* = 1.18, *p* = .237).

#### iii) Fatigue and quality of life

Finally, the three groups differed on insomnia (*H* = 6.66, *p* = .036), fatigue (*H* = 6.45, *p* = .040), vitality (*H* = 6.50, *p* = .039), and emotional wellbeing (*H* = 9.18, *p* = .010). The other effects were not significant (*p* > .05 for all comparisons).

The mild patients reported more fatigue than the severe patients (*z* = -2.57, *p* = .010), while there were no differences between either the mild and moderate patients (*z* = -0.71, *p* = .481) or both moderate and severe patients (*z* = -1.52, *p* = .130). Pairwise comparisons did not reach significance after Bonferroni correction (mild vs. moderate: *z* = -1.99, *p* = .046; mild vs. severe: *z* = -2.28, *p* = .023; moderate vs. severe: *z* = -0.71, *p* = .481).

Conversely, severe patients reported more vitality, emotional wellbeing and social function than mild patients (vitality: *z* = 2.65, *p* = .008; wellbeing: *z* = 2.97, *p* = .003; social function: *z* = 2.99, *p* = .002). Pairwise comparisons between severe and moderate patients did not reach significance after Bonferroni correction (wellbeing: *z* = 2.01, *p* = .044; vitality: *z* = 1.06, *p* = .290; social function: *z* = 1.66, *p* = .097), nor did those between mild and moderate patients (wellbeing: *z* = 0.68, *p* = .494; vitality: *z* = 1.12, *p* = .263; social function: *z* = 1.06, *p* = .290) (see Table 4).

### Relationships between neuropsychological deficits, psychiatric symptoms, and other secondary variables

The third aim was to examine whether the presence of long-term neuropsychological deficits was correlated with psychiatric symptoms and/or other clinically relevant variables.

The results of the multiple regression performed on each cognitive variable are set out in Table 5. Interestingly, apathy, depression, anxiety, emotion regulation, emotion contagion, stress, PTSD, dissociative disorders, anosmia and diabetes all proved to be variables of interest when it came to explaining the neuropsychological sequelae. Therefore, both psychiatric and nonpsychiatric data correlated with neuropsychological deficits across the three groups. There were at least three patterns of results, depending on the neuropsychological domain: i) patterns in which neuropsychological sequelae did not correlate with any psychiatric variables, but did with other clinical variables, such as visuospatial long-term episodic memory (delayed recall of Rey-Osterrieth complex figure); ii) patterns in which neuropsychological sequelae correlated with both psychiatric and clinical variables, such as the object and action naming task scores (language); and iii) patterns in which neuropsychological sequelae only correlated with psychiatric variables, such as categorical verbal fluency. There was also a fourth possible pattern where the neuropsychological sequelae correlated neither with psychiatric variables nor with clinical ones, such as the score on the object decision task (object perception).

**Table 5.**
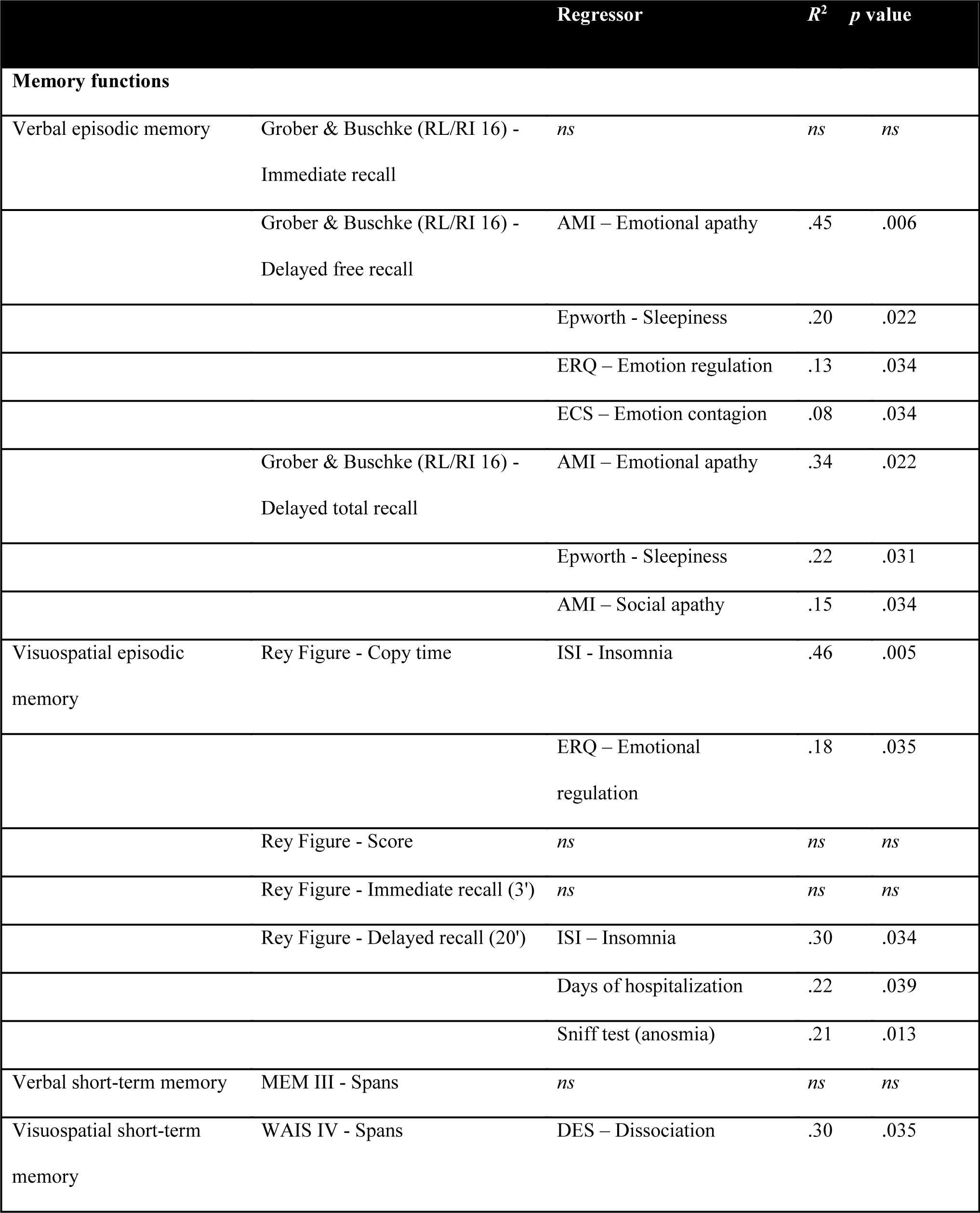
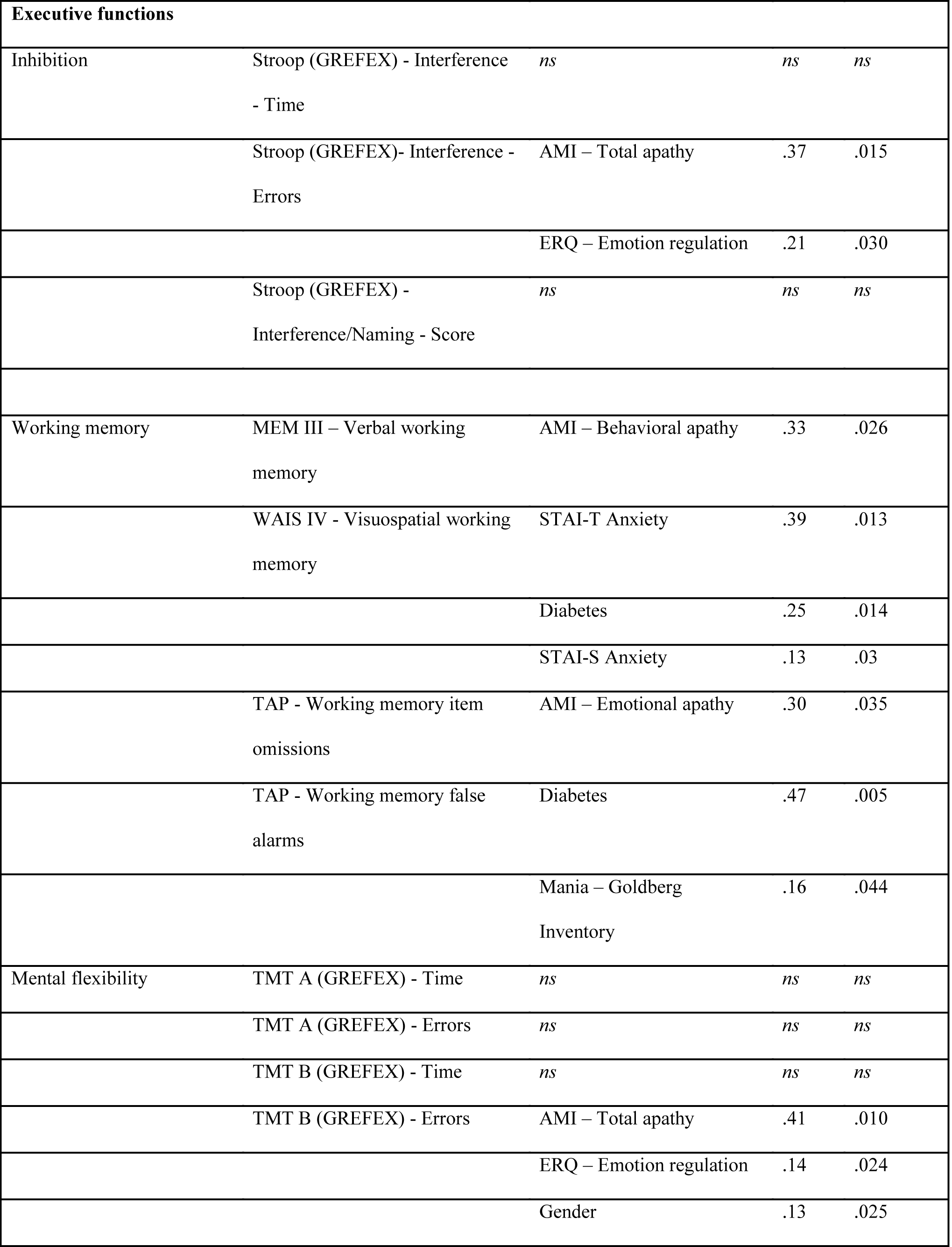
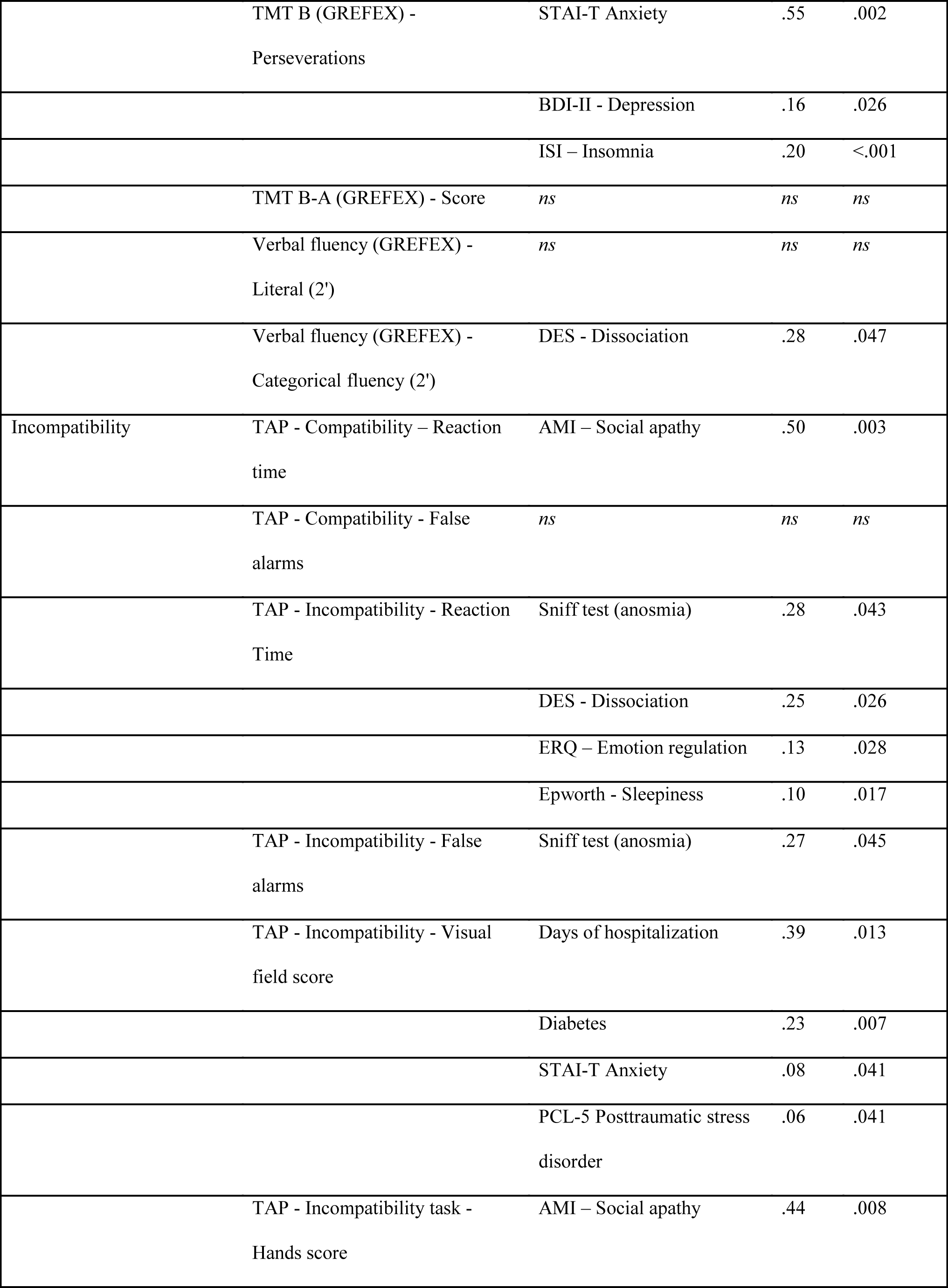
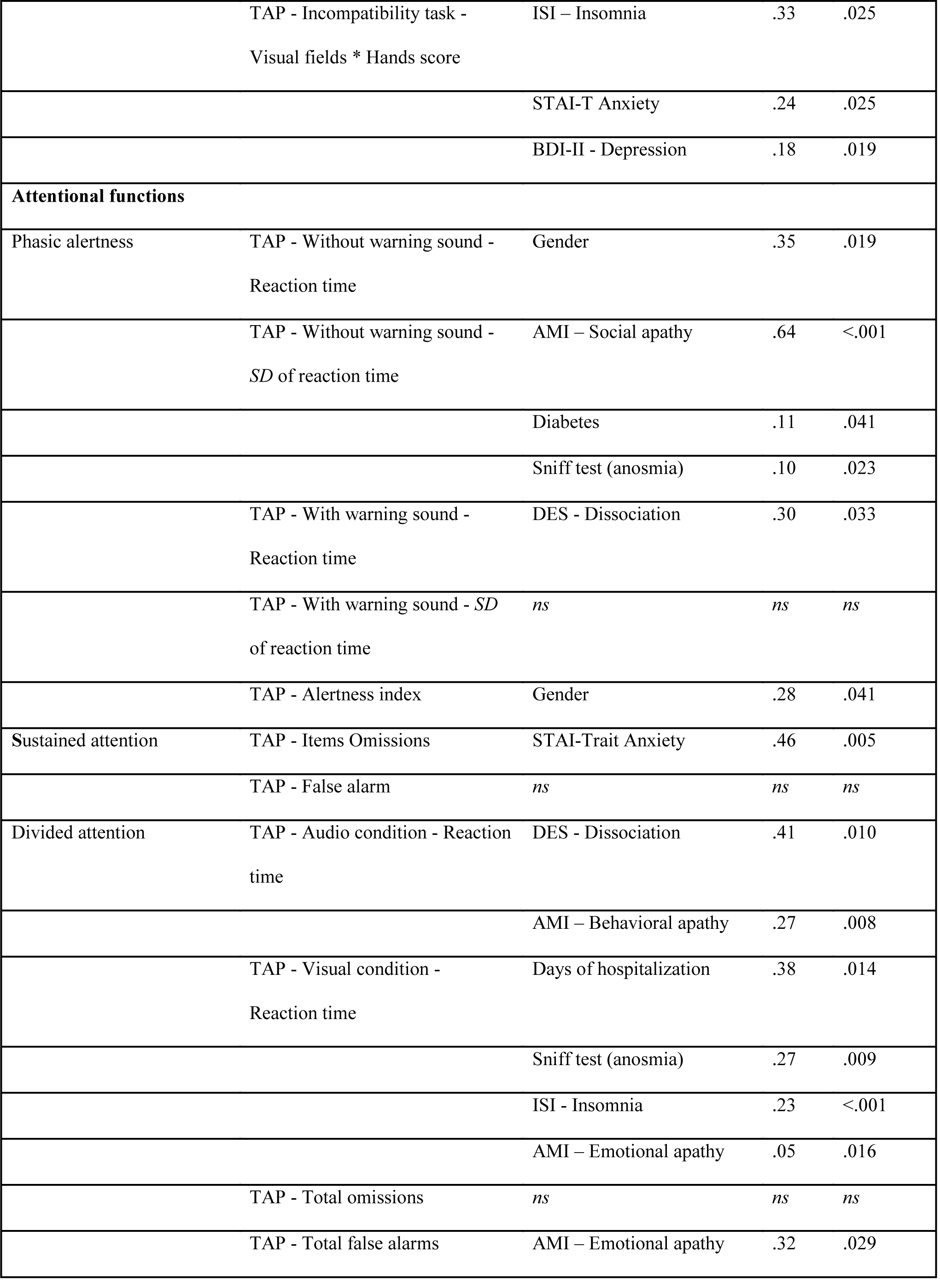
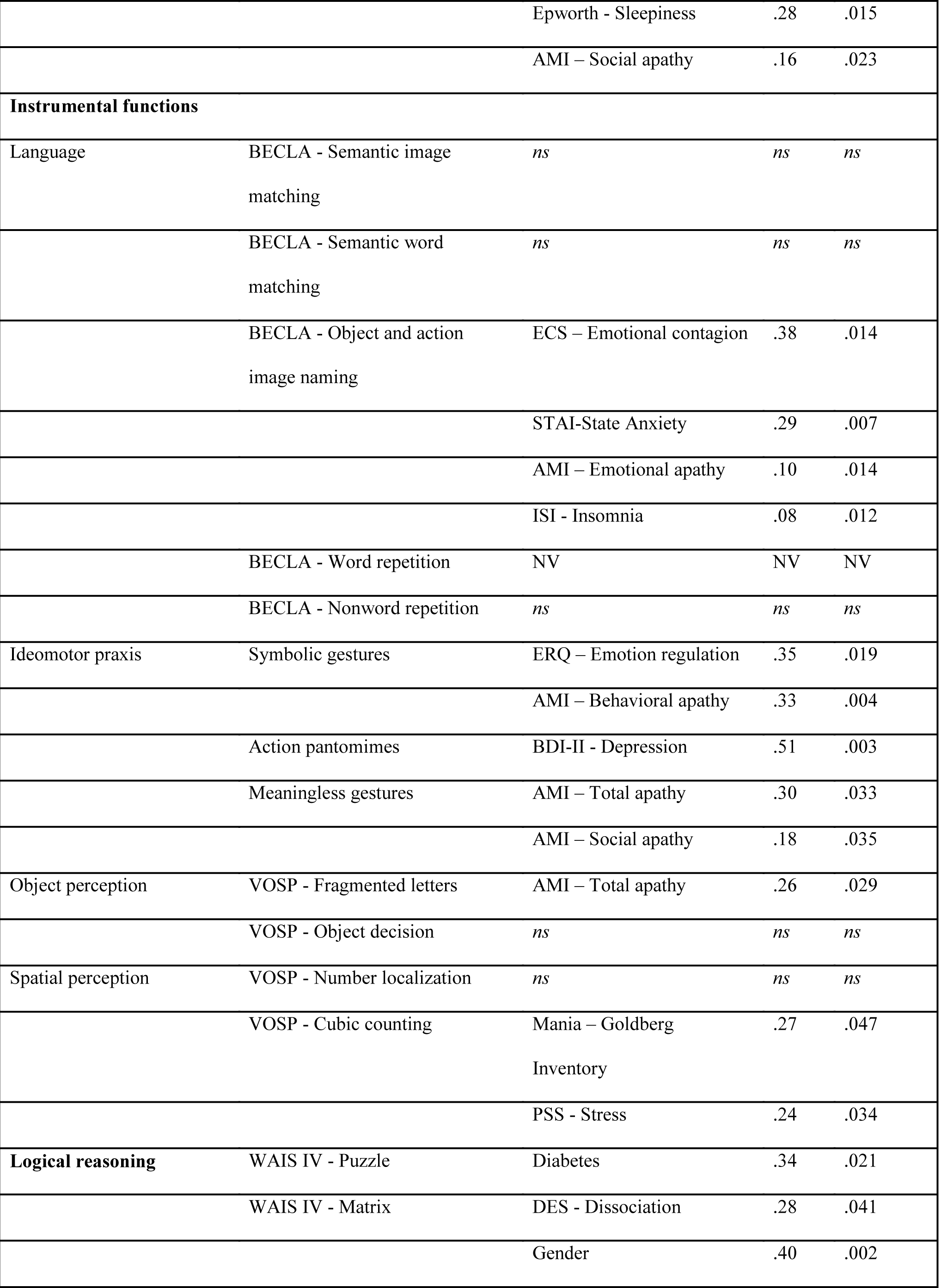
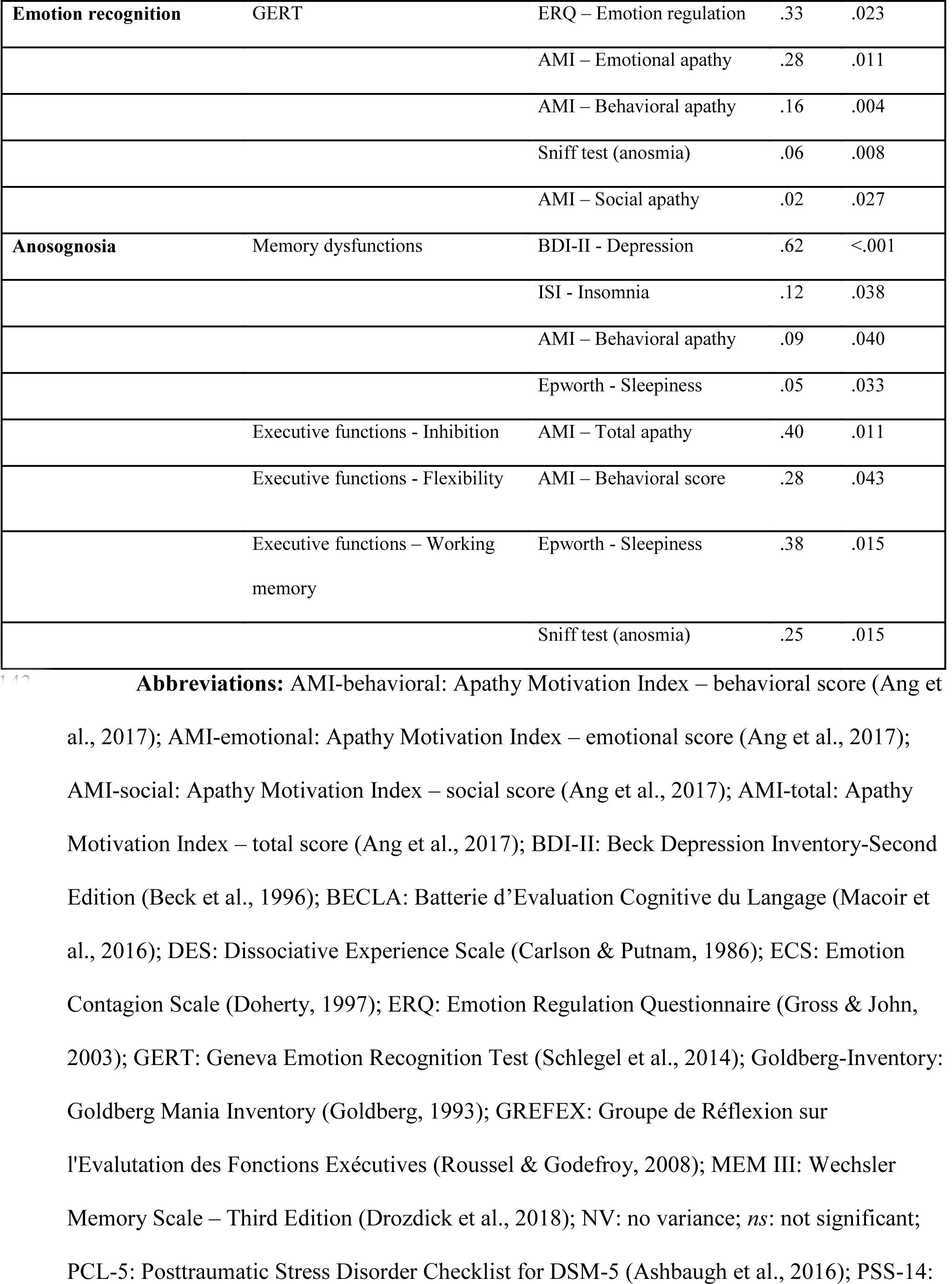
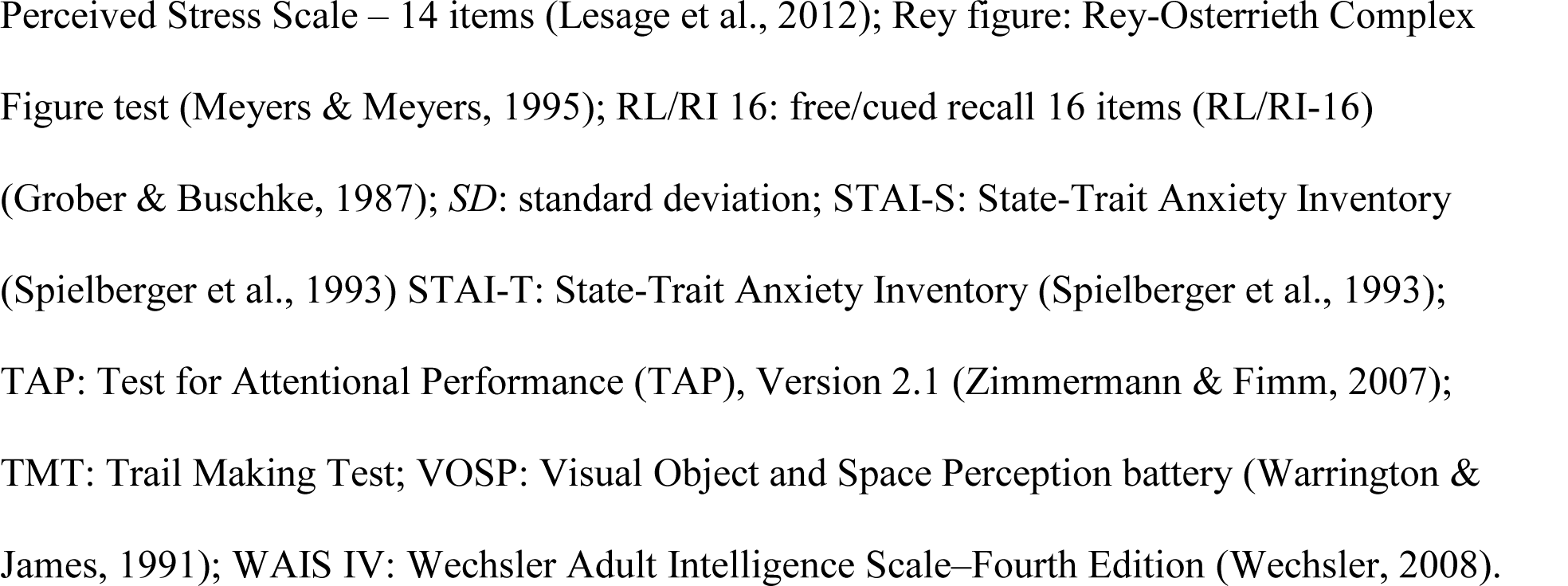
Multiple regression results for each of the neuropsychological variables

To reduce the dimensionality of the dataset, we computed a PCA. We selected the first three orthogonal components accounting for 43.67% of the total variance. The first component, accounting for 26.75% of the total variance, was difficult to interpret in terms of underlying cognitive processes, as it included language (semantic word and image matching), executive functions (mental flexibility), verbal episodic memory, and emotion recognition.

Interestingly, these happened to be precisely the variables on which the three groups differed significantly (see Section *“Neuropsychological and psychiatric symptoms as a function of disease severity”*). We therefore labeled this component *respiratory disease severity*. The second component (9.79% of total variance) was labeled *attention and anosognosia*, as it included alertness, divided attention, and anosognosia for executive dysfunction. The third component (7.15% of total variance) was labeled *instrumental functions*, as it included language, visual perception, and ideomotor praxis.

For the respiratory disease severity component, the best fit was achieved with emotional apathy (*R*^2^ = .28, *p* = .007), stress (*R*^2^ = .19, *p* = .013), and anosmia (*R*^2^ = .11, *p* = .03). For the attention and anosognosia component, the multiple regression was not significant (*p* > .1). For the instrumental functions component, the best fit was achieved with anosmia (*R*^2^ = .23, *p* = .04), mania (*R*^2^ = .23, *p* = .006), and social apathy (*R*^2^ = .17, *p* = .004).

## DISCUSSION

Even though there is growing evidence that SARS-CoV-2 can cause brain damage in the long term, with an impact on cognition even in its mild and moderate forms, to date, the occurrence and nature of such sequelae, the impact of respiratory disease severity in the acute phase, and the relationship between these impairments and psychiatric disorders triggered or exacerbated by the pandemic have not been studied in detail within a single sample of patients. In addition, areas such as instrumental functions (ideomotor praxis, visual perception, or language), cognitive complaints, anosognosia, and emotion recognition following SARS-CoV-2 have yet to be explored. Finally, the relevant medical events have not been controlled in studies published thus far. The present study used a robust, psychometrically validated methodology and a stringent approach to the normative data of neuropsychological tests (excluding borderline scores from the prevalence calculation). We included patients with no history of cancer or neurological and developmental disorders, and no active psychiatric disorders before SARS-CoV-2 infection, and divided them into mild, moderate and severe groups, according to the respiratory severity of the disease during its acute phase.

The present study therefore improves our understanding of what we can call *neurological long COVID*, highlighting three main patterns of results. First, important prevalence of patients *across the three groups* performed below the normality threshold in all domains of cognition (except ideomotor praxis) 6-9 months post-infection with SARS-CoV-2. The prevalence of psychiatric symptoms, regardless of disease severity during the acute phase, was also high, and individuals in all three groups exhibited depressive symptoms, anxiety, mania, apathy, stress, PTSD and dissociative disorders, as well as reporting insomnia, fatigue and pathological somnolence. Regarding olfaction, 33.33% of the mild group, 73.33% of the moderate group, and 46.66% of the severe group were still hyposmic 6-9 months following infection, and 13.33% of the severe group were still anosmic. Second, despite the presence of common cognitive deficits across the three groups, some domains of cognition and mood were differentially impacted by the severity of respiratory disease during the acute phase: the severe group performed more poorly than the mild group on long-term episodic memory, and also exhibited more anosognosia for memory dysfunction. The mild group was more depressed, stressed and anxious, and reported more cognitive complaints. Finally, the moderate group recognized multimodal emotions less well than the mild group. All of this had a substantial impact on patients’ quality of life. Third, as predicted, neuropsychological deficits correlated with psychiatric disorders such as depressive symptoms, stress and mania, but not all of the variance was explained by psychiatric symptoms or transdiagnostic syndrome (Husain & Roiser, 2018). A large proportion of the variance was explained by other clinical variables. For instance, the long-term episodic memory deficits displayed by the severe group were positively correlated with emotional apathy, their anosognosia for memory dysfunction was correlated with depression, and their diminished emotion recognition, shared by the moderate group, was positively correlated with hyposmia and/or anosmia.

The present study had several limitations that need to be acknowledged and addressed before we can draw any inferences from our results. The first drawback was a possible recruitment bias. By enrolling volunteers, we may have selected the most severe cases in the mild group (who were interested in the study because of their cognitive complaints), while we may not have recruited the most cognitively affected in the severe group, because they were too disabled to join the study. Second, we had greater proportions of men and diabetics in the severe group. These factors may have had an influence on the cognitive deficits observed in this group, as diabetes is known to impact cognition (McCrimmon, Ryan, & Frier, 2012), and gender on depression (Spagnolo, Manson, & Joffe, 2020), with a greater prevalence in women (Mazza et al., 2020). That said, although the proportion of women was higher for both the mild and moderate groups, the mean depression scores by gender in the mild (women: 13.50 ± 9.10; men: 12.57 ± 8.52) and moderate (women: 6.11 ± 5.25; men: 13.33 ± 11.25) groups did not indicate a greater proportion of women with depressive symptoms. Third, stroke is more prevalent in patients after a severe SARS-CoV-2 infection (Merkler et al., 2020; Nannoni et al., 2020), and may have gone unseen during the acute phase. In our study, no patient had any central neurological deficit excluding major stroke, but minor stroke cannot be ruled out. Two patients in the severe group reported mild signs of peripheral neuropathy, which may have been due to their diabetes and not a direct consequence of the SARS-CoV-2 infection, while one patient in the severe group had an unstable gait. Fourth, the absence of a control group prevented us from observing a possible general effect of the pandemic and the resulting public health measures on mental health. In the present study, the analyses of prevalence were based on standardized normative data, allowing us to run comparisons with the normal population.

The tests were chosen carefully for their psychometric validity, with adequate sensitivity and specificity. It is interesting to note that in a recent study, our multimodal emotion recognition task (GERT) was administered to 469 participants during the pandemic (Schlegel, von Gugelberg, Makowski, Gubler, & Troche, 2021) but the authors failed to find a reduction in performances compared with validation studies (Schlegel et al., 2014), reinforcing the hypothesis that our results reflected a specific effect of the infection and not just the public health context. Fifth and last, it is important to note the relatively small number of participants, which prevented us from considering more covariates. Nevertheless, the power analysis, based on a previous study of the neurocognitive effects of SARS-CoV-2, did allow us to estimate the necessary sample size.

The present results demonstrate that cognitive deficits can be observed 6-9 months post-SARS-CoV-2 infection, regardless of the severity of the disease in the acute phase. They corroborate previous observations for the executive, attentional and memory domains, and go one step further, with exhaustive neuropsychological and psychiatric assessments demonstrating impairments in other previously unexplored cognitive and psychiatric domains. Impairments were evident not only in the severe patient group, but also in the moderate and mild groups. These deficits had an impact on quality of life, notably in the mild patients, as evidenced by our results. These findings could be of great importance in understanding the long-term damage and consequences of SARS-CoV-2 infection on cognition and mental health. The relatively high prevalence of certain cognitive and psychiatric disorders, regardless of the severity of the disease in the acute phase, suggests that long-term patient management following SARS-CoV-2 infection may need to be adapted. Importantly, the etiology of these disorders needs to be established, in order to provide people experiencing these long-term sequelae with the best possible care. One potential explanation for these effects, based on observational studies of the psychiatric impact of the pandemic in the general population (Bäuerle et al., 2020), is that these cognitive deficits result from a stressful or traumatic context. In this case, specific interventions on certain psychiatric variables could considerably reduce their long-term impact on cognition and improve daily functioning.

Nevertheless, the present results do not exclude the hypothesis of direct damage of brain networks by SARS-CoV-2 and its neurotropism, as well as indirect neurobiological effects, which could lead to both to psychiatric and neurological disorders. COVID-19 may induce CNS disturbance, and four main pathogenic mechanisms may act in combination: i) direct viral encephalitis, ii) systemic inflammation, iii) peripheral organ dysfunction (liver, kidney, lung), and iv) cerebrovascular changes (Iadecola, Anrather, & Kamel, 2020). At this stage, it is difficult to determine whether the cognitive deficits can be regarded as a marker of brain damage, and/or should be linked to psychiatric variables that may themselves result directly from infection with SARS-CoV-2 or else be triggered by the stressful nature of the general pandemic and the individual experience of the disease.

Second, this study highlighted the presence of differential cognitive and psychiatric profiles at 6-9 months post-infection as a function of the respiratory severity of the SARS-CoV-2 infection in the acute phase. This suggests the existence of different clinical phenotypes. In the identification/discrimination of these phenotypes, different cognitive variables seem to be of interest, starting with cognitive complaints and anosognosia. While the severe patients exhibited anosognosia for their memory dysfunction and greater long-term verbal memory impairment than the mild patients did, the latter had more cognitive complaints. This fits in well with the observations of Almeria et al. (2020), who found that the patients with the most serious cognitive complaints did not have significantly more neuropsychological impairments. In this sense, the tendency of the severe patients to report greater wellbeing in quality-of-life assessments, together with the lack of awareness of their cognitive difficulties, may be a clinical characteristic to bear in mind when interviewing this type of patient. The present results in the domain of emotion recognition and episodic memory are also highly relevant to the current debate on the neurotropism of SARS-CoV-2.

One of the main hypotheses regarding the pathways of direct attack of the CNS assumes olfactory transmucosal invasion by the virus (Meinhardt et al., 2021). This hypothesis appears to be supported by our results. It is worth noting that episodic memory and emotion recognition were identified in a PCA as variables that explained most of the variance of our data, and this first component was significantly correlated with hyposmia/anosmia, in addition to stress and emotional apathy. Interestingly, a recent ^18^F-FDG PET study demonstrated hypometabolism at about 8 weeks post-infection in brain regions common to emotion and olfaction in patients with SARS-CoV-2 (Guedj et al., 2020). Moreover, the literature suggests that the viral load was probably greater in our severe group (Fajnzylber et al., 2020; Magleby et al., 2020), which may have contributed to stronger effects on olfaction and emotion recognition. This pathway could also partially explain the psychiatric results via disruption of the limbic network, including subcortical regions (Lane, 2008), by SARS-CoV-2.

Our third level of analysis enabled us to go further in characterizing the hypothesized clinical phenotypes. Quantified results pointed to the presence of at least three profiles (patient clusters), corroborating the clinical impressions we had when interviewing and assessing the patients for this study. Patients with the first (neurological) profile were typically aged about 55 years, mostly men, of average educational level, and a small proportion of them had a history of diabetes, cardiovascular disease, or sleep apnea syndrome. At the cognitive level, these patients displayed long-term memory, executive and language disorders. They had more severe anosognosia for their memory difficulties. Nearly all of them reported sleep disorders and emotional apathy. Patients with the second (psychiatric) profile were aged about 45-50 years, and there were equal numbers of men and women. No significant medical antecedents were noted, and the majority of them had had mild or moderate respiratory disease. At the cognitive level, they displayed executive and attentional dysfunctions, which could influence other cognitive domains (e.g., memory recall strategies). At the psychiatric level, they had high scores for depressive symptoms, anxiety, insomnia and stress, and more sporadically exhibited PTSD and dissociative disorders. Our results also indicated the presence of a third (mixed) profile combining the symptoms and clinical characteristics of the two previously described profiles.

## CONCLUSION

This study unambiguously demonstrates the presence of long-term neuropsychological sequelae following SARS-CoV-2 infection, regardless of the severity of the respiratory disease in the acute phase. Some of the cognitive deficits could be explained by psychiatric variables, emphasizing the importance of considering a broad range of psychiatric symptoms. However, not all neuropsychological sequelae could be explained by these variables. The presence of correlations between olfaction, emotion recognition and episodic memory, which share common functional and anatomical substrates, reinforces the hypothesis that the virus targets the CNS (and notably the limbic system). Finally, the data support the notion of different clinical phenotypes, paving the way for clinical guidelines and recommendations for the management of long-term neurological impairment following SARS-CoV-2 infection.

## Data Availability

Data will be avaible on Yareta (University of Geneva). The repository is under progress.

## ACKNOWLEDGEMENTS

The present research was supported by the Swiss National Science Foundation (SNSF) within the framework of the COVID-19 National Research Program (NRP 78; grant no. 407840_198438, RNP 78) to JAP (PI) and FA (Co-PI). The funders had no role in data collection, discussion of content, preparation of the manuscript, or decision to publish. We would like to thank the patients for contributing their time to this study.

## SUPPLEMENTARY INDEX

Supplementary Index 1. Raw scores (cognitive tests; psychiatric questionnaires included in the principal component analysis).

Grober & Buschke (RL/RI 16) - Immediate recall

Grober & Buschke (RL/RI 16) - Delayed free recall

Grober & Buschke (RL/RI 16) - Delayed total recall

MEM III - Spans

MEM III - Working verbal memory

WAIS IV - Spans

WAIS IV - Visuospatial working memory

Rey Figure – Copy score

Rey Figure - Immediate recall (3’)

Rey Figure - Delayed recall (20’)

Stroop (GREFEX)- Interference - Errors

Stroop (GREFEX)- Interference/naming - Score

TMT B-A (GREFEX) - Score

Verbal fluency (GREFEX) - Literal (2’)

Verbal fluency (GREFEX) - Categorical (2’)

TAP phasic alertness - Without warning sound - Reaction time

TAP phasic alertness - With warning sound - Reaction time

TAP phasic alertness - Alertness index

TAP sustained attention - Item omissions

TAP sustained attention - False alarm

TAP divided attention - Audio condition - Reaction time

TAP divided attention - Visual condition - Reaction time

TAP divided attention - Total omissions

TAP divided attention - Total false alarms

TAP Incompatibility task - Visual fields * Hands score

BECLA - Semantic image matching

BECLA - Semantic word matching

BECLA - Object and action image naming

BECLA - Word repetition

BECLA - Nonword repetition

Evaluation of gestural praxis - Symbolic gestures

Evaluation of gestural praxis - Action pantomimes

Evaluation of gestural praxis - Meaningless gestures

VOSP - Fragmented letters

VOSP - Object decision

VOSP - Number localization

VOSP - Cubic counting

WAIS IV - Puzzle

WAIS IV - Matrix

GERT – Emotion recognition task

Anosognosia - Memory functions

Anosognosia - Executive functions - Working memory

Anosognosia - Executive functions - Inhibition

Anosognosia - Executive functions - Flexibility

